# Discrete-Event Simulation Modeling Framework for Cancer Interventions and Population Health in R (DESCIPHR): An Open-Source Pipeline

**DOI:** 10.1101/2025.05.12.25327470

**Authors:** Selina Pi, Carolyn M. Rutter, Carlos Pineda-Antunez, Jonathan H. Chen, Jeremy D. Goldhaber-Fiebert, Fernando Alarid-Escudero

**Affiliations:** Department of Biomedical Data Science, School of Medicine, Stanford University, 300 Pasteur, Edwards, Floor 3, Palo Alto, CA 94304; Hutch Institute for Cancer Outcomes Research, Biostatistics Program, Public Health Sciences Division, Fred Hutch Cancer Center, 1100 Fairview Ave, Seattle, WA 98109; The Comparative Health Outcomes, Policy, and Economics (CHOICE) Institute, University of Washington, 1959 NE Pacific St, Seattle, WA 98195; Stanford Center for Biomedical Informatics Research, Stanford University, 3180 Porter Dr, Palo Alto, CA 94304; Stanford Clinical Excellence Research Center, Stanford University, 453 Quarry Rd, Palo Alto, CA 94304; Division of Hospital Medicine, Stanford University, 453 Quarry Rd, Palo Alto, CA 94304; Department of Health Policy, School of Medicine, Stanford University, 615 Crothers Way, Stanford, CA 94305; Center for Health Policy, Freeman-Spogli Institute for International Studies, Stanford University, 615 Crothers Way, Stanford, CA 94305

**Keywords:** decision-analytic modeling, discrete-event simulation, microsimulation, cancer, screening, Bayesian calibration

## Abstract

Simulation models inform health policy decisions by integrating data from multiple sources and forecasting outcomes when there is a lack of comprehensive evidence from empirical studies. Such models have long supported health policy for cancer, the first or second leading cause of death in over 100 countries. Discrete-event simulation (DES) and Bayesian calibration have gained traction in the field of Decision Science because they enable flexible modeling of complex health conditions and produce estimates of model parameters that reflect real-world disease epidemiology and data uncertainty given model constraints. This uncertainty is then propagated to model-generated outputs, enabling decision makers to assess confidence in recommendations and estimate the value of collecting additional information. However, there is limited end-to-end guidance on structuring a DES model for cancer progression, estimating its parameters using Bayesian calibration, and applying the calibration outputs to policy evaluation. To fill this gap, we introduce the **DES** Modeling Framework for **C**ancer **I**nterventions and **P**opulation **H**ealth in **R** (DESCIPHR), an open-source codebase integrating a flexible DES model for the natural history of cancer, Bayesian calibration for parameter estimation, and an example application of screening strategy evaluation. To illustrate the framework, we apply DESCIPHR to calibrate bladder and colorectal cancer models to real-world cancer registry targets. We also introduce an automated method for generating data-informed parameter prior distributions and increase the functionality of a neural network emulator-based Bayesian calibration algorithm. We anticipate that the adaptable DESCIPHR modeling template will facilitate the construction of future decision models evaluating the risks and benefits of health interventions.

**Key points for decision makers:**

1. For simulation models to be useful for decision-making, they should accurately reproduce real-world outcomes and their uncertainty.
2. The DESCIPHR framework and code repository address a gap in open-source resources to fit an individual-level model for cancer progression to real-world data and forecast the impact of cancer screening interventions while accounting for data uncertainty.
3. The codebase is designed to be highly adaptable for researchers who wish to apply DESCIPHR for economic evaluation or for studying methodological questions.

## 1 Introduction

Decision-analytic models inform policy decisions by integrating available evidence in a mathematical framework to forecast outcomes under alternative intervention strategies and potential counterfactual scenarios [1]. In particular, microsimulation models have had concrete effects on cancer screening policy, for instance informing the United States (US) Preventive Services Task Force’s (USPSTF) breast cancer screening guidelines starting from 2009 [2] and a reduction in the recommended start age for US colorectal cancer (CRC) screening in 2021 [3, 4]. Compared to discrete-time microsimulation methods that model the state of a system at fixed time steps, discrete-event simulation (DES) models gain efficiency by only focusing on the time points of events at which a system changes [5] and more easily incorporate competing risks and time-varying patient characteristics [6]. These features have made DES a compelling alternative for model-based economic evaluations and operations research in healthcare settings, leading to its application for modeling health systems, disease progression, screening, and patient behavior [7]. Microsimulation models often contain deep, or unobservable, parameters that must be estimated through calibration. Calibration seeks to identify parameters such that the model reproduces real-world clinical, biological, or epidemiological outcomes, such as disease incidence and subtype prevalence [8, 9]. Bayesian calibration methods, in particular, have become widely used in cancer modeling [10–17] and other health applications, such as modeling infectious diseases [18, 19] or smoking behavior [20], to output parameter distributions that capture the measurement uncertainty of the real-world quantities used as calibration targets. Though Bayesian calibration can be computationally demanding, advances in model emulator methods, statistical approximations, and computing resource utilization have made them faster to implement and deploy [13, 21].

Though published methods, codebases, and best practice guidelines provide some instruction for implementing DES and Bayesian calibration separately, there is limited integrated and publicly available guidance with tutorial code on implementing and calibrating DES cancer models. Fewer than 10% of publications with healthcare DES models release open-source versions of their models [22], and open-source DES tools, such as the simmer R package [23] and the SimPy [24] and Ciw Python libraries [25], are more suited for operations questions than for disease natural history modeling. The steep learning curve of DES with existing bespoke open-source models has prompted the release of a tutorial on cost-effectiveness analyses of screening in R using a simplified DES-based cancer model [26], but the tutorial does not cover parameter estimation and calibration. Open-source cancer simulation models, such as the Colon Modeling Open Simulation Tool (CMOST) [27] and Microsimulation Lung Cancer (MILC) model [10, 28] are implemented with discrete-time methods or restrictive distributional assumptions and more limited calibration techniques. Open-source programs for Bayesian calibration of disease models are limited in scope, stopping short of integrating the required data pre-processing to prepare calibration targets for the model and applying the calibration outputs to decision analyses [11, 13–15, 21, 29].

To facilitate the development of individual-level models for cancer policy evaluation, we introduce the **DES** Modeling Framework for **C**ancer **I**nterventions and **P**opulation **H**ealth in **R** (DESCIPHR) with an open-source codebase for 1) structuring a DES model for cancer natural history, 2) estimating model parameters with state-of-the-art Bayesian calibration methods, and 3) applying the calibrated model for policy analyses accounting for parameter uncertainty, illustrated with a screening evaluation example.

The pipeline also augments Bayesian Calibration using Artificial Neural Networks (BayCANN) [21] to include hyperparameter tuning and multiple output types for constrained calibration targets. The model is built in R because of its accessibility and well-maintained and documented packages for statistical analysis and calibration, making it the most commonly used free and open-source programming language for healthcare DES models [22, 30–32]. The structure of the code repository follows the Decision Analysis in R for Technologies in Health (DARTH) framework for decision modeling, designed to facilitate model transparency and adaptability [32]. Section 2.1 introduces the DES cancer natural history model. Sections 3 and 4 explain parameter estimation of the model using modern Bayesian calibration methods and screening policy evaluation using the parameters’ joint posterior distribution, respectively. In Section 5, we apply DESCIPHR to bladder and colorectal cancer to demonstrate how it can be adapted to real-world cancer registry targets and used to simulate two different types of cancers: one with precancerous lesions, colorectal, and one without, bladder. We provide the code for the entire pipeline and cancer site-specific analyses on GitHub at https://github.com/sjpi22/tutorial cancer modeling des.

## 2 Cancer natural history model

Microsimulation modeling for policy analysis requires simulating a disease under natural history, that is, in the absence of intervention, as a baseline against which the impacts of interventions are compared [33]. There are multiple approaches to modeling the natural history of cancer, ranging from highly detailed cellular-level simulations [34, 35] to population-level cohort and microsimulation models [28, 36–38]. In practice, the key is to build the simplest model with just enough structural detail to appropriately evaluate the intervention(s) of interest and maintain face validity [33, 39]. Accordingly, the modeling process should begin by articulating the cancer control interventions to evaluate (e.g., primary prevention, screening, active surveillance, treatment), identifying the aspects of disease natural history that are influenced by the intervention (for example, the transition from undetected to screen-detected disease, removal of precursor lesions, or post-diagnosis survival), and specifying any other features that differentiate the intervention’s effect in the model. In this section, we describe the structure, DES implementation, and summary outcomes of a model capturing the sequence of landmark events in cancer natural history needed to evaluate screening. In Appendix A, we provide guidance on implementing the model and extending it to other applications, such as cancer surveillance and treatment evaluation.

### 2.1 Disease states and transitions

We model the natural history of cancer by splitting the carcinogenic process into health states reflecting disease onset, progression, diagnosis, and death, consistent with the structure used in numerous general and site-specific cancer simulation models [12, 27, 28, 37, 40–45]. Relevant health states and possible transitions between states are illustrated in the model diagram in Figure 1. In the model, patients are assumed to be born into a healthy state, labeled as state *H* in Figure 1. If the natural history of a cancer site includes precursor lesions, such as adenomas for CRC [46–48] or cervical intraepithelial neoplasia (CIN) for cervical cancer [49], the onset of a lesion causes patients to transition to the precancerous lesion state, labeled as state *L*. For simplicity, the current implementation of the model assumes a single lesion type and no lesion regression, although guidance on incorporating additional cancer subtypes and lesion growth patterns is provided in Appendix A.4. Additional lesions may develop over the course of the individual’s life. At the first conversion of any lesion to detectable cancer, patients enter the preclinical, or pre-detection, cancer stage (state *P*). The model can also account for a direct transition from the healthy to the preclinical cancer state in the absence of precancerous lesions.

**Fig. 1:**
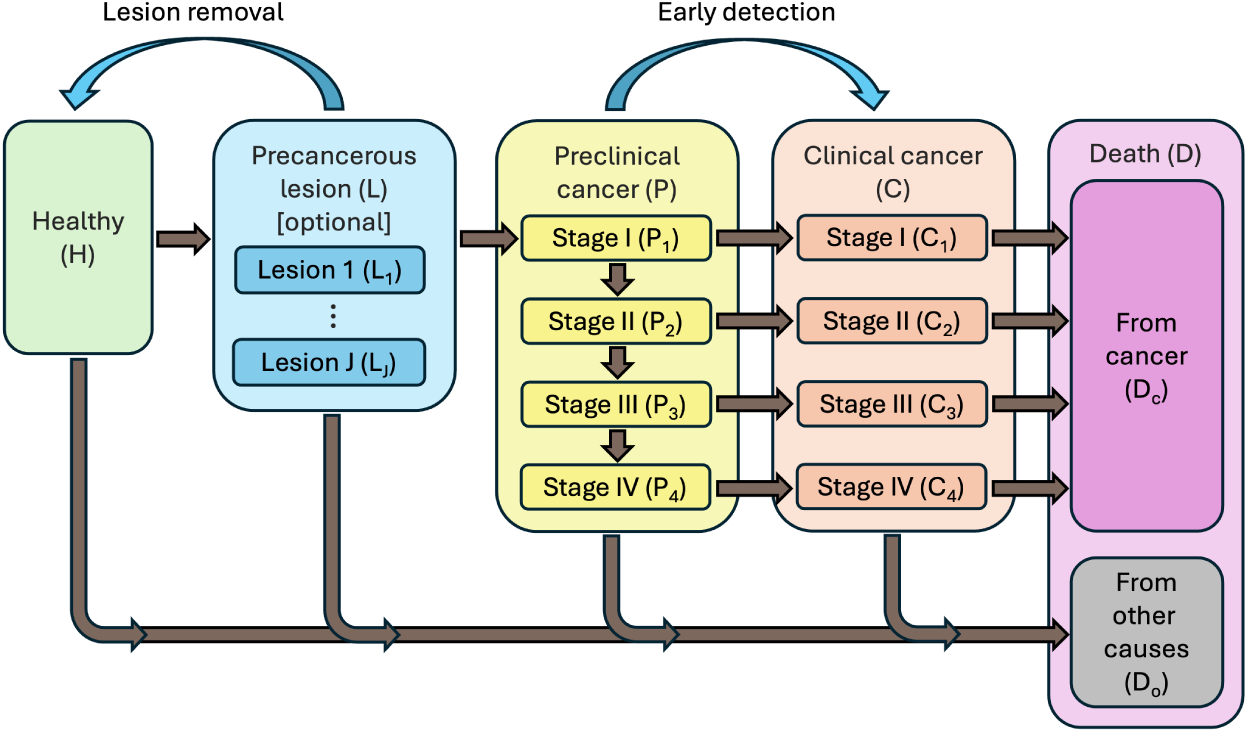
Cancer natural history model schema with effects of screening in blue arched arrows

Within the preclinical cancer state, patients advance through progressive stages of disease until symptom-based diagnosis, upon which they transition to the clinical cancer state (state *C*). There are multiple approaches to modeling cancer progression, such as simulating the sojourn time and tumor size before assigning the stage at detection based on size [12] or modeling the hazard of stage progression and detection as proportional to tumor volume [50]. In our implementation, for each stage before the final stage, we sample the time to progression to the next stage and the time to symptomatic detection within the stage. The stage and time of diagnosis are determined by the first stage during which the time to detection occurs earlier than the time to progression, or the time to detection within the last stage if detection has not occurred before then. Individuals who develop clinical cancer face a risk of dying from cancer, thus transitioning to the state *D_c_*, based on a stage- and disease-specific mortality rate derived from relative survival data from cancer registries. Cancer treatment effects are not explicitly modeled, but we assume that stage-specific relative survival distributions account for the survival associated with current technology. All individuals are at risk of death from other causes (state *D_o_*), following an age-specific mortality rate derived from life tables and independent of the cancer pathway. The time of death is determined by the earlier of death from cancer and death from other causes, with the death state denoted as *D*. Guidance on customizing the model structure, for example, by adding an infection state preceding the precursor lesion state, is provided in Appendix A.4.

### 2.2 DES implementation

We simulate the mathematical model described in Section 2.1 using a DES approach [51] by defining the events as transitions between states. When generating patient trajectories using DES, the time from entering state *i* to entering a consecutive state *j* is characterized as a random variable *T_i,j_* that follows a time-to-event (TTE) distribution with probability density function *f_i,j_*, cumulative distribution function (CDF) *F_i,j_*, and hazard function 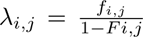. The hazard can be a function of the time *t* spent in state *i*, age *a*, time period *p*, and other demographic and risk factors *x*, *i.e.*, *λ_i,j_*(*t, a, p, x*), to account for heterogeneity related to population dynamics, genetic and environmental influences, treatment effects, and age-related biological responses.

Individual transition times and state characteristics can be generated using inverse transform sampling given the corresponding CDF, meaning that for each individual *n*, a value 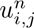 s drawn from a standard uniform distribution and used to calculate the time 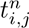 at which *F_i,j_* would equal 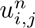 :

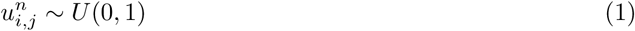

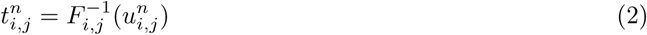

TTE variables may also be sampled using intensity-based methods [52]. For the results to be reproducible, a fixed random seed should be set before running an analysis based on random sampling.

After the times between consecutive states are sampled, the time from entry into the health state *i* to entry into a non-consecutive future health state *k* can be calculated as the sum of times within consecutive states from *i* to *k*. The model generates a matrix with a row for each individual in the simulated cohort and a column for each simulated patient characteristic, TTE variable, or health state characteristic. For models that include the precancerous lesion state, an additional lesion-level event matrix is generated with a row for each lesion that develops during a patient’s lifetime and columns for the times of lesion onset and conversion to preclinical cancer, as well as other modeled lesion characteristics. More details on the implementation of the probability distributions and sampling procedures are provided in Appendix A.

### 2.3 Data inputs and parameters

While some distributions characterizing the model transitions and state characteristics may be obtained from published literature or empirically estimated with individual-level data using well-established survival analysis methods, others are unobserved but may be estimated through calibration to real-world data sources, a process described in Section 3. In the former category, the time to death from other causes is often modeled empirically using background mortality rates derived from actuarial life tables, as done in several established cancer models [12, 38, 41], with adjustments for disease-specific rates if they substantially impact all-cause mortality [53, 54]. In addition, the time from cancer diagnosis to death from cancer is commonly sampled from empirical relative survival distributions by stage at diagnosis, which are reported by the US National Cancer Institute’s (NCI) Surveillance, Epidemiology, and End Results (SEER) program [55].

Conversely, no data exist to directly inform most pre-diagnosis distributions, such as the time to cancer onset, dwell time within preclinical stages, and rate of precancerous lesion development, which need to be estimated via calibration. These unobserved transitions are modeled by assigning a distributional form and then calibrating the distribution parameters to selected real-world outcomes, known as calibration targets. Epidemiological outcomes commonly used as calibration targets include the prevalence of precancerous lesions and incidentally detected cancer in screening or autopsy studies [11, 13, 15, 56, 57], the incidence and stage distribution of clinical cancer from cancer registry data in the absence of screening [2, 11, 15, 56, 58], the proportions of the population with different subtypes of precancerous lesions or cancer [57], and the multiplicity and size distribution of lesions [12, 15]. For a cancer natural history model, age-specific preclinical cancer prevalence and symptomatic cancer incidence are important for informing the transition rates to cancer onset and symptom-based detection by age, while the stage distribution informs the distribution of disease progression at diagnosis, which predicts survival outcomes. Targets for lesion characteristics inform tumor growth rates and heterogeneity in cancer pathways. Outcomes from screening and surveillance trials [45, 59] have also been used for calibration.

Choosing the distributional form for unobserved transitions is a somewhat subjective process but may be guided by the calibration targets. Since most cancer types display an increasing risk with age [60], a parametric distribution with an accelerating hazard, such as a Weibull distribution, can be a good starting point to model the time of precancerous lesion or cancer onset [37, 61], though sigmoidal functions have also been used to reflect tapering incidence at older ages [27]. Due to their simplicity, exponential distributions, which have a constant transition rate, are also commonly used for unobserved distributions [11, 41, 44, 62], although they may not adequately describe transitions with a nonzero mode or long tail [63]. Misspecification of the distributional form may bias estimates of outcomes [37], so conducting model validation and sensitivity analyses is encouraged to examine which distributional assumptions produce a reasonable model fit after calibration (see Section 3.4).

### 2.4 Summary outcomes

After generating event data using an individual-level simulation model with a given set of parameters, the data must often be aggregated into summary statistics reflecting population-level outcomes for various purposes, including model calibration, validation, and policy evaluation. Both calibration and validation of the natural history model, described in Section 3, involve comparing modeled outcomes to epidemiological endpoints estimated from empirical data, as discussed in Section 2.3. For prevalence, incidence, stage distribution, multiplicity, and survival targets, we simulate a population representative of the real-world sample from which the endpoint is derived and calculate the corresponding epidemiological outcomes using the equations described by [64]. We may also estimate intermediate outcomes of cancer natural history that are often not observed in practice, including the mean sojourn time of cancer and the mean dwell time of precancerous lesions, according to the formulas in [64]. For decision analysis, tradeoffs associated with screening may be calculated in terms of quality-adjusted or absolute life years gained (LYG) and the economic or resource burden from additional testing compared to no screening. Average LYG is calculated by summing total life-years, optionally among individuals meeting specific criteria (e.g., alive and cancer-free at a given age), dividing by the cohort size to obtain per capita estimates, repeating the calculation under screening (see Section 4.2), and computing the difference. Incremental testing burden can be calculated similarly. Because life trajectories in the microsimulation model are generated through random sampling, the simulated outcomes exhibit stochastic variation that depends on the simulated population size. In Appendix A.3, we provide an approach to determine the cohort size accounting for this variation.

## 3 Parameter estimation with Bayesian calibration

As discussed in Section 2.2, disease models may include deep parameters that are not estimable using individual-level data but may be inferred through calibration, which involves adjusting the parameters so that the model outputs, when summarized according to Section 2.4, are consistent with real-world calibration targets [10]. Various methods exist to search the parameter space for optimal-fitting parameters, including manual tuning, empirical search, directed search, and Bayesian techniques [33]. Whereas directed search algorithms output a single parameter set, we focus on Bayesian calibration methods because they combine prior knowledge about the parameters of interest with the uncertainty associated with the calibration targets to output a distribution of parameters reflecting their joint uncertainty.

In Bayesian calibration, the modeler specifies prior distributions for unknown model parameters and measures the goodness-of-fit for the targets given the parameters using a loss function, such as a likelihood. The output is the joint posterior distribution for the parameters, which can then be sampled to simulate distributions for model outcomes of interest. For each outcome, we can generate base case estimates from the expected value and 95% posterior model-prediction intervals (PI) from the 2.5th and 97.5th percentiles of the simulated outcomes from the posterior distribution. An early limitation of Bayesian calibration was the computational intensity required to simulate large numbers of model samples in sequence to achieve accurate estimates of the joint posteriors [10, 65, 66]. However, advances in computational capacity and statistical approximations have made Bayesian methods increasingly feasible.

Approximate Bayesian computation (ABC) refers to a family of likelihood-free methods that infer model parameters by simulating outputs and assessing how closely they match observed or target data, thereby approximating the likelihood [67]. Incremental mixture ABC (IMABC) leverages ABC in an initial rejection-sampling step and then performs adaptive sampling of regions consistent with targets; the adaptive sampling is similar to incremental mixture importance sampling (IMIS) and allows IMABC to overcome inefficiencies of ABC in searching high-dimensional parameter spaces and calibrating to multiple targets [13]. Since the most time-consuming aspect of Bayesian calibration is simulating the model with different parameter samples, emulators that quickly and accurately map model inputs to desired outputs, such as the Bayesian Calibration using Artificial Neural Networks (BayCANN) algorithm, could drastically reduce runtime [15, 21]. IMABC and some steps in BayCANN can be parallelized to optimize computational resources and speed up processing.

### 3.1 Choice of priors

Assigning prior distributions to calibrated model parameters may be guided by meta-analysis of evidence, expert opinion, biological knowledge, and common sense to reflect the state of knowledge before model fitting [65, 66, 68]. When there is little evidence to inform the parameters, one can perform empirical calibration, such as Nelder-Mead optimization followed by a grid search, to set bounds for mutually independent, uniformly distributed diffuse priors [10, 29]. In Appendix B, we also describe a target-informed approach to derive priors for the parameters underlying the time from birth to disease onset. Prior to calibration, to ensure the model can produce outputs in the ranges of the calibration targets, we perform a model coverage analysis by sampling from the joint prior distribution, plotting the model outputs, and demonstrating that the model-predicted outputs have a wider range than the 95% confidence intervals of the calibration targets. For BayCANN, the coverage analysis can be conducted using the parameter and output set generated to train the neural network emulator.

### 3.2 IMABC

IMABC improves on earlier Bayesian calibration algorithms, such as Markov Chain Monte Carlo (MCMC) and ABC, due to its ability to handle high-dimensional parameter spaces and its compatibility with parallel processing. The initial rejection-based sampling step draws *N_o_* parameter sets from the prior distribution using Latin hypercube sampling (LHS) and accepts sets that produce model outputs within a user-specified tolerance threshold from the targets [13]. Parameter sets are ranked using a distance-based measure [69]. At each iteration of the subsequent updating phase, the algorithm samples from a mixture of multivariate normal distributions centered at the *N* ^(^*^c^*^)^ highest ranking parameter sets and keeps the sets whose outputs fall within incrementally narrowing bounds around the targets. The algorithm stops when either the number of accepted parameter sets within the final target bounds reaches a user-specified value *N_post_* or the maximum number of iterations is reached. It is recommended to use a large *N_o_* (for example, 1000 samples per calibrated parameter) and wide initial tolerance intervals to increase the algorithm’s likelihood of identifying acceptable parameter sets in the first step [13]. Additional guidance for choosing the values of these arguments is provided in [13] and the IMABC GitHub documentation. We used the IMABC R package to implement Bayesian calibration with IMABC. The output of the calibration is a set of accepted parameters with weights that account for the adaptive sampling. A weighted sample of the posterior distribution is used to generate decision-analytic outcomes.

### 3.3 BayCANN

A metamodel, or emulator, is a function that approximates a more computationally intensive simulation model by predicting model outputs based on the same inputs [70]. BayCANN uses an artificial neural network (ANN) as an emulator to map parameters of the cancer natural history model to summary statistics corresponding to the calibration targets [21] and has been applied to emulate other models, including a seasonal influenza model [71] and the CISNET CRC models [14]. Popular in machine learning due to their accuracy in modeling nonlinear interactions between input variables, ANNs are a type of regression consisting of layers of interconnected nodes that resemble the interconnected neurons of the brain [72]. BayCANN includes sample generation, emulator training, and parameter estimation steps as illustrated in Figure 2. We increase its functionality from the original implementation with two modifications: incorporating hyperparameter tuning during ANN training to increase emulator accuracy and enabling multiple output layers in the ANN, allowing it to handle different activation functions and directly integrate constraints on the outputs.

**Fig. 2:**
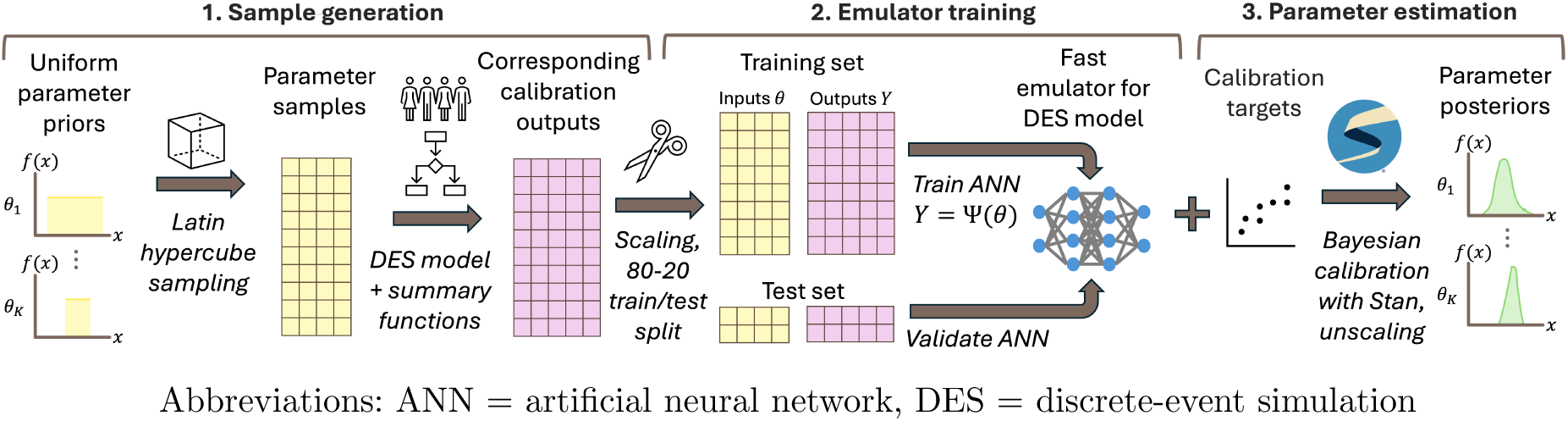
Steps of BayCANN

**Fig. 3:**
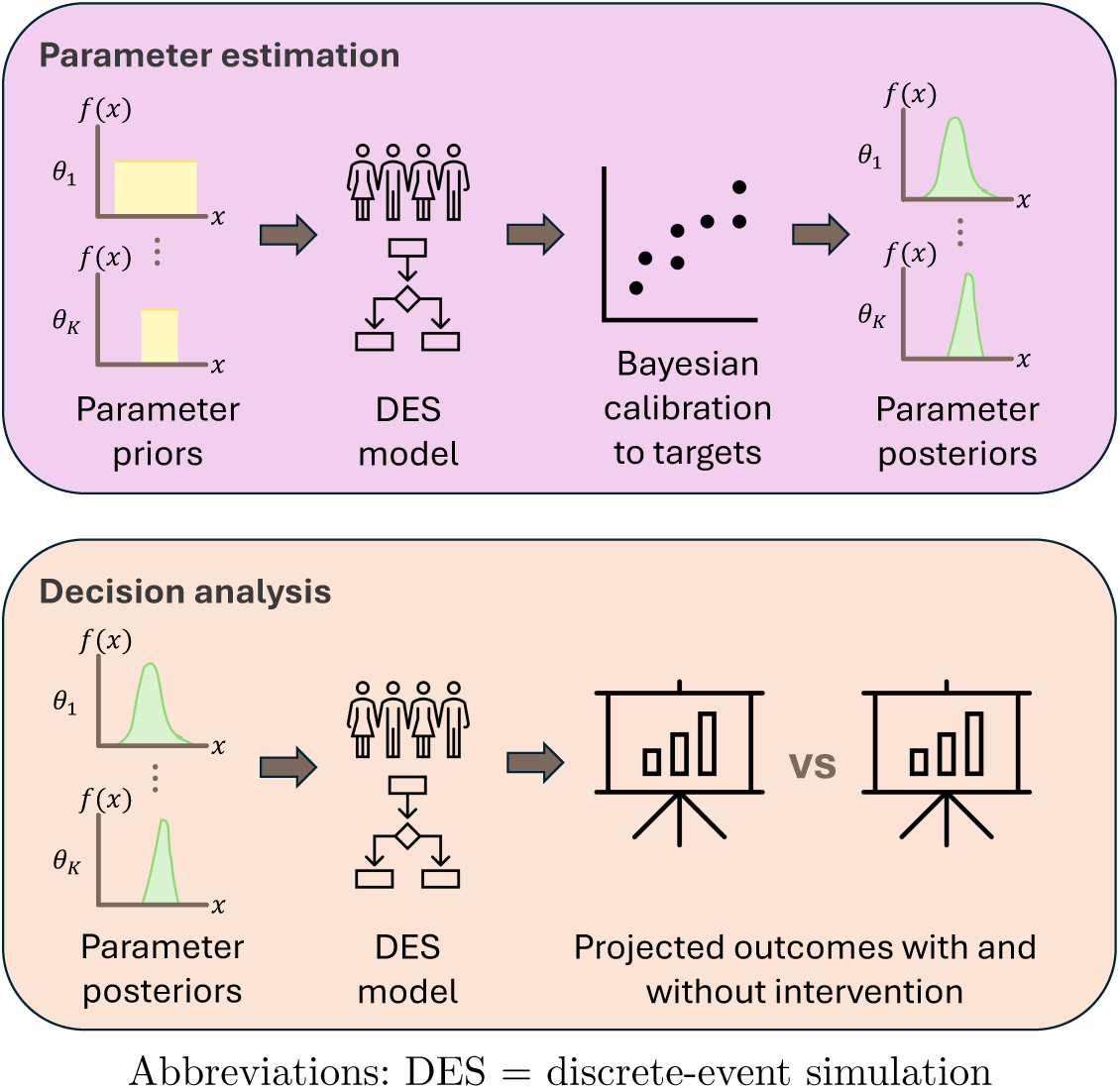
Diagram of DESCIPHR modeling framework

#### 3.3.1 Sample generation

To create a sample for ANN fitting and validation, we draw parameter sets from the prior distributions using LHS; simulate a cohort of life trajectories for each set; and calculate the simulated outcomes corresponding to the calibration targets for each cohort. The sets of parameters and outcomes are divided for training and testing, often with a ratio of 80-20, and standardized to ensure stable gradients during ANN training.

#### 3.3.2 Emulator training

We fit the ANN by minimizing a loss function, such as the mean squared error or binary cross-entropy, between the ANN’s predicted outputs and the cancer model outcomes for the training set. The training set may be further split into training and validation sets for hyperparameter tuning, which is the process of testing various values of ANN hyperparameters to identify the configuration that yields the most accurate ANN. Hyperparameters include the number of layers, number of nodes per layer, type of activation function, inclusion of dropout layers, and dropout rate. We may assign groups of outputs with different output activation functions, losses, evaluation metrics, and contribution weights in the total loss. For the output layer, sigmoid activation is consistent with targets such as prevalence and standardized outputs that range from 0 to 1, softmax activation for groups of variables that must sum to 1, exponential or rectified linear unit (ReLU) activation for nonnegative outputs, and linear activation for outputs with no range constraints. We implement the ANN and hyperparameter tuning using the keras3 and tfruns R packages. The ANN can be validated by calculating the loss between the predicted and true outputs in the held-out test set or plotting the ANN-predicted outputs against the model outputs. Lines of points along the diagonals indicate that the emulator accurately predicts the model outputs.

#### 3.3.3 Parameter estimation

The hyperparameters, weights, and biases of the final keras3 ANN are extracted and input into an ANN coded in the probabilistic programming language Stan via the rstan interface [73]. Using the Hamiltonian Monte Carlo algorithm, Stan generates a collection of parameters that approximates the joint posterior distribution and that can be uniformly sampled to produce new model outputs. To assess the quality of the Stan model fit, we check the mixing of the chains and the R-hat convergence diagnostic. The posteriors are returned to the original scale of the parameters.

### 3.4 Internal validation

Validating the calibration results establishes trust in the model’s predictions and involves evaluating the consistency of the model with the calibration targets (internal validation), other models (comparative model validation), or data held out from the calibration process (external validation) [33, 74, 75]. For internal validation, we visually examine whether the model outputs produced by sampling from the joint parameter posterior distribution (see Section 4.1) replicate the targets and their uncertainty bounds [75]. The IMABC algorithm automatically generates the calibration outputs associated with the posteriors. For BayCANN, the user may choose to sample from the posterior as described in Section 4.1 and simulate the calibration outputs and decision outcomes together for efficiency. For a more detailed discussion of the implementation and reporting of comparative and external validation, which are stronger forms of validation, we refer to [74–76].

### 3.5 Nonidentifiability

If available calibration targets are insufficient to inform all calibrated parameters, these parameters may be nonidentifiable, meaning that there are multiple parameter sets that fit equally well with the calibration targets but may have different policy implications [77]. For instance, a natural history characterized by a slow onset time but fast sojourn time may fit observed cancer incidence data just as well as one with a fast onset time and slow sojourn time, but screening intervals would need to be shorter in the former case to intercept cancers at the same rate. In such cases, it is important to transparently report how the model transitions and parameter prior distributions are constructed and to simulate outcomes from the entire posterior distribution of the parameters, ensuring that policy analyses reflect their uncertainty. Parameter nonidentifiability can be addressed by using informative priors, but this approach requires justification, as informative priors may bias the posterior [68, 78].

## 4 Model-based screening policy evaluation

We illustrate an application of the modeling and calibration pipeline by sampling from the parameter posterior distribution and other uncertain model inputs to estimate the effects of screening. In this section, we first provide general guidance on how to apply the calibration results to policy evaluation and then describe how screening interventions interact with the natural history model.

### 4.1 Sampling from the posterior

The outputs obtained from Bayesian calibration with IMABC and BayCANN are a random sample of parameter sets from the joint posterior distribution of the model parameters. For each parameter set in the sample, we can estimate the impact of a particular intervention by simulating a cohort of individual trajectories with the natural history model, sampling from distributions that capture the uncertainty of quantities related to the intervention (for example, the sensitivity of a screening test or the hazard ratio of a cancer treatment), updating the life trajectories in response to applying the intervention to the cohort, and comparing outcomes of interest between the scenarios with and without the intervention. The distribution of outputs associated with the parameter posterior sample and other uncertain quantities constitutes a probabilistic sensitivity analysis (PSA) of the impact of the intervention. This procedure can be repeated for different interventions to compare outcomes, determine the optimal policy recommendations, and assess the uncertainty in the recommendations and estimates.

### 4.2 Modeling screening

Screening may intervene in the disease process in two ways: it can prevent cancer by removing precursor lesions, thereby interrupting their progression to cancer, as in the case of CRC or cervical cancer; or it may modify the disease process through the early detection of cancer, potentially at a more treatable stage, as in the case of prostate and breast cancer. However, mass screening in individuals without disease is associated with clinical and economic burdens, including the cascade of care triggered by false positive results [79, 80]. To evaluate the effects of screening, a cohort of individual trajectories is first simulated without it (i.e., under natural history). Next, screening regimens are applied to generate counterfactual trajectories, from which we estimate the effects of screening on cancer-related outcomes. We consider the LYG and test burden compared to the natural history scenario, which are two of many outcomes, including quality of life, adverse events, healthcare costs, and equity, that policymakers may consider when deciding whether to adopt a new screening intervention. Costs can also be assigned to each test to capture the economic burden of screening.

We define a screening regimen by its starting age, stopping age, screening interval, and test characteristics. Individuals who are alive without diagnosed cancer by the screening starting age will undergo screening until the earliest of the screening stopping age, the diagnosis of cancer, or death. Test characteristics required for modeling include the sensitivity *p*, which is the probability of a positive result among individuals with disease, and specificity *q*, or the probability of a negative result for an individual without disease. If any precancerous lesions are removed at a particular screening event, the time to preclinical cancer onset is recalculated as the earliest conversion time of any remaining lesions over the individual’s lifetime. Before applying the next test, the time to death from cancer is recalculated by adding the original times from preclinical cancer onset to clinical cancer and then to death from cancer, assuming independence between the age of preclinical cancer onset and the time from onset to symptom-based diagnosis. Under the same assumption, the stage of symptom-based diagnosis remains unchanged. Meanwhile, the confirmation of asymptomatic cancer causes the patient to transition immediately to the detected cancer state, and the time to death from cancer is resampled based on the stage at diagnosis. The time to death is then recalculated as the minimum time to death from either cancer or other causes but restricted to occur no earlier than in the no-screening scenario.

## 5 Application to bladder and colorectal cancer

To illustrate the modeling and calibration approaches, we apply DESCIPHR to two cancer types that differ in their natural history structure: bladder cancer, which has no precursor pathway, and CRC, which does.

### 5.1 Bladder cancer

Bladder cancer is associated with a lifetime risk of 3.3% in men and 1.0% in women [55]. It is typically diagnosed after blood is detected in a routine urine test, leading to a cystoscopy to visualize the bladder wall [81]. Currently, there is no population-based bladder cancer screening program in the US, but there is interest in researching and modeling the potential benefits of early detection and active surveillance [82, 83].

#### 5.1.1 Data

For the background mortality distribution informing the time to death from non-cancer causes, we use the 1933 life table for the overall US population from the Human Mortality Database [84]. For the time from diagnosis to death from cancer, we use SEER relative survival rates from 2000 to 2021 [55]. Since relative survival is only reported up to 10 years from diagnosis, we extrapolate beyond 10 years by fitting a concave B-spline to the cumulative hazard of cancer-specific mortality. For calibration targets, we use SEER data on age-specific incidence in 5-year intervals from 0 to 85 and aggregated for the population 85 and older, and stage distribution (localized, regional, and distant) of bladder cancer from 1989 to 1993, the earliest period reported in the SEER archives [85]. We exclude unstaged cancer and increase the percentages of the other categories proportionally so that they sum to 1.

#### 5.1.2 Natural history model

We model the natural history of bladder cancer using the model described in Section 2.1 without the precursor lesion state between the healthy and preclinical cancer states. That is, healthy individuals transition directly to preclinical cancer. The transitions between states are parameterized by the distributions listed in Table 1. Although the natural history of bladder cancer includes carcinoma in situ as a stage preceding the localized stage, we only model progression starting from the localized stage due to data limitations. The natural history model ultimately contains 8 parameters that must be calibrated to the incidence and stage distribution targets.

**Table 1:**
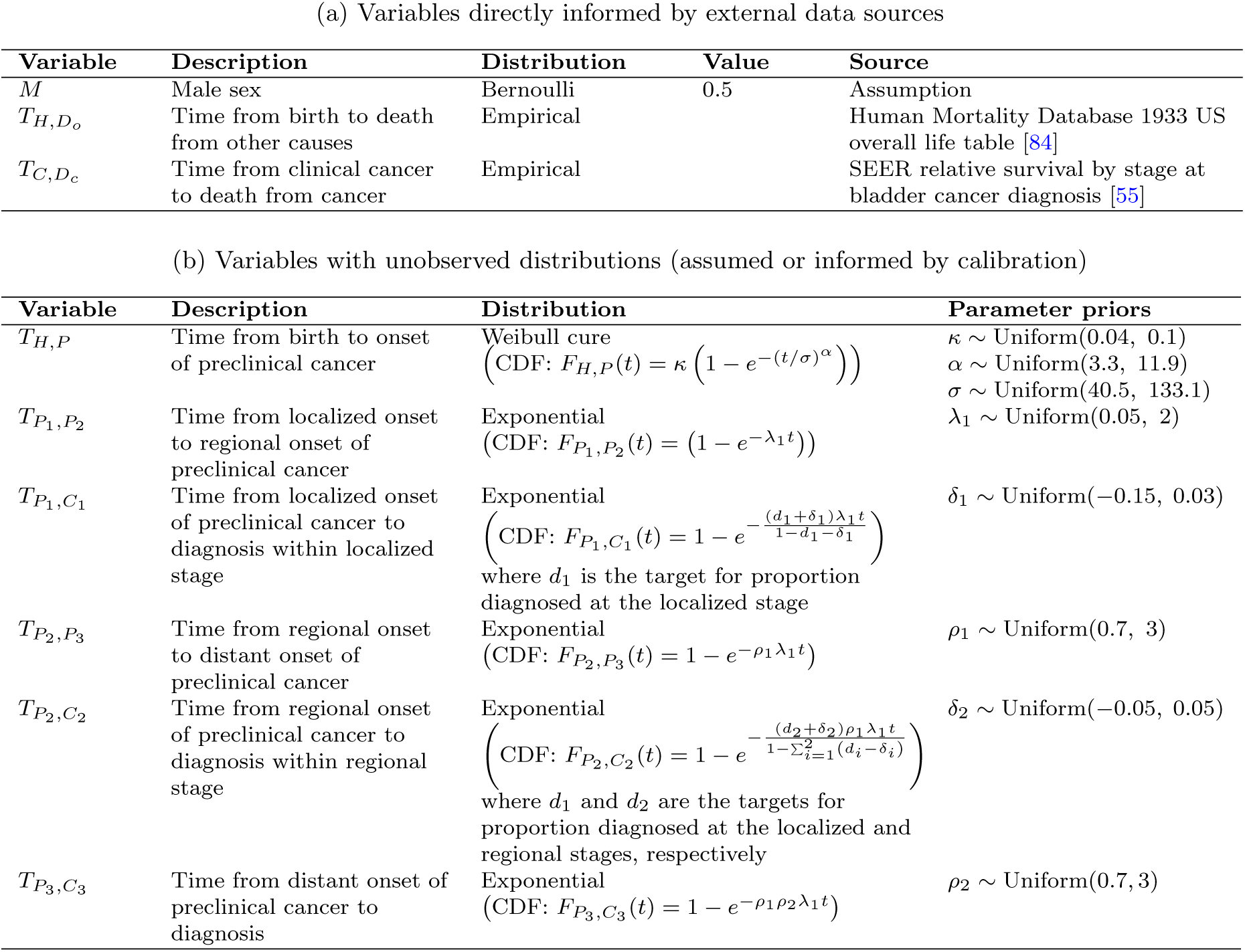
Bladder cancer model parameters and distributions.

#### 5.1.3 Parameter estimation with IMABC

To determine the prior bounds for the calibrated parameters, we apply the procedure in Appendix B to estimate a range of parameters for the time to preclinical cancer onset and use a wide range of plausible priors for the other parameters. Since data on the age-specific prevalence of preclinical bladder cancer was unavailable, we use the ratio of incidental to known bladder cancer diagnoses in autopsy data from [86] and SEER cancer incidence to estimate age-specific prevalence for calculating the prior bounds. We use the (1 − 0.0001) × 100% confidence interval of the calibration targets as the final bounds for the simulated outputs, as the sample size of the SEER registry leads to narrow bounds. Following the procedure in Appendix A.3 and multiplying the target cohort size by 2, we calculate that a cohort size of 1,500,000 would achieve a Monte Carlo error within these bounds.

For the initial bounds of the calibration targets, we expand the final bounds by three times their difference from the target value. We run IMABC with an initial sample of 4000 parameter sets (500 samples per calibrated parameter) drawn from the prior distributions using LHS to identify sets associated with model outputs within the initial bounds. After the initial step, IMABC continues to run by sampling 50 parameter sets per center from a multivariate normal mixture distribution with 5 centers for up to 1200 iterations or until the posterior distribution achieves an effective sample size of 1000. The algorithm ran for 12 hours on 16 cores on a high-performance computing cluster, ending with 395 in-range parameter sets. For internal validation, the 95% model-predicted posterior ranges of outputs are shown in gray against the final bounds of the calibration targets in red in Figure 4, showing good overlap.

**Fig. 4:**
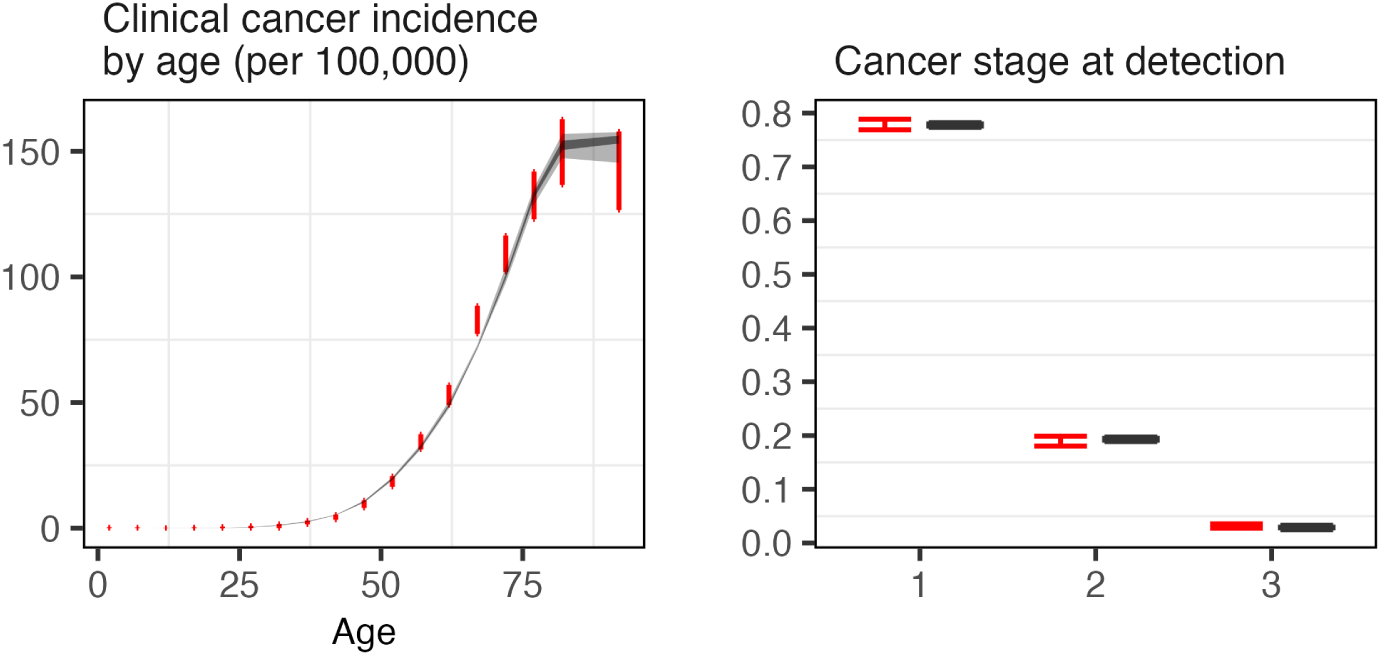
Internal validation of calibrated bladder cancer model with model outputs in gray and (1 − 0.0001) × 100% CIs of calibration targets in red listed in Table 2. The natural history model ultimately contains 13 parameters that must be calibrated to adenoma prevalence and colorectal cancer incidence and stage distribution targets.

**Table 2:**
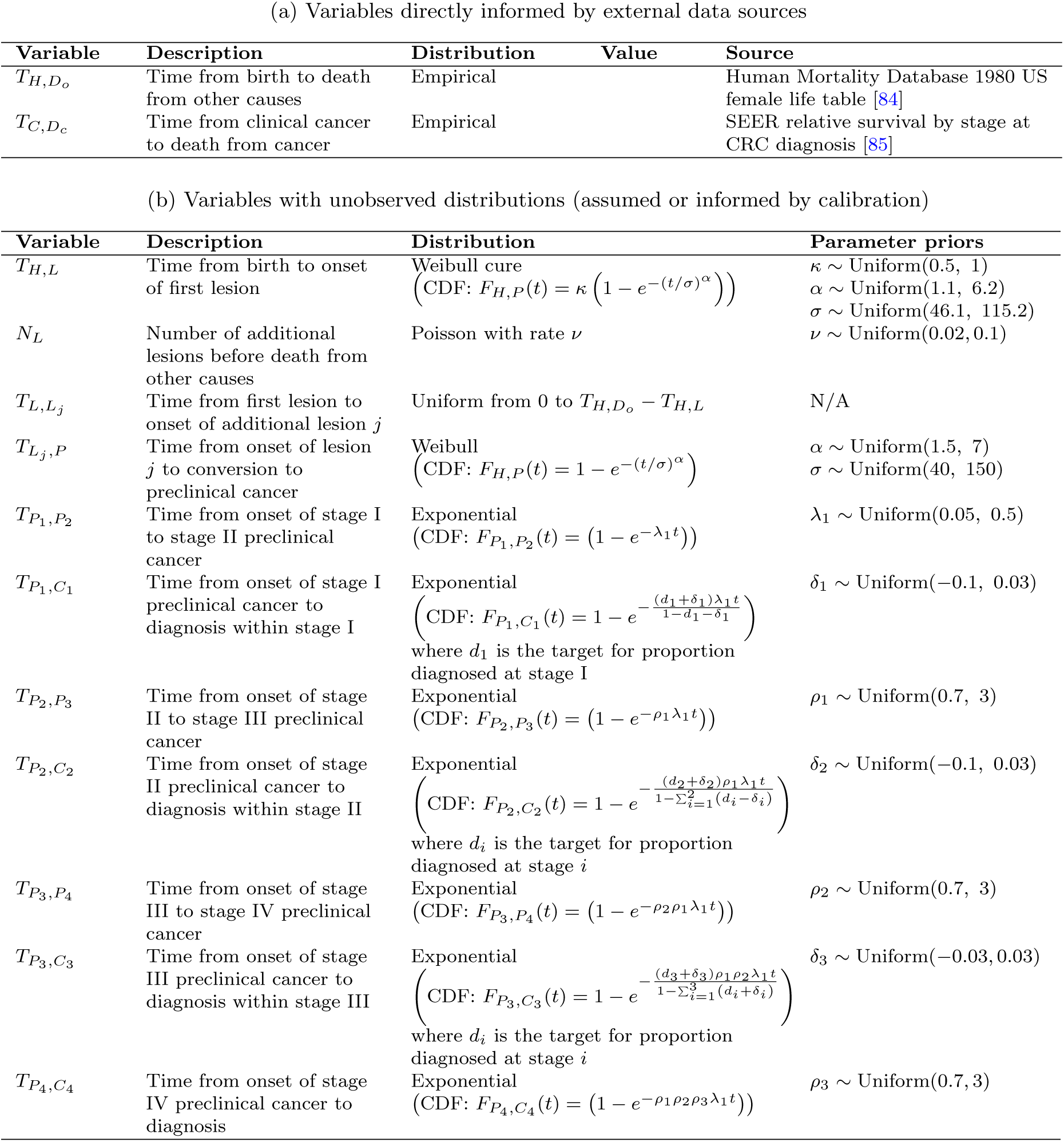
Colorectal cancer model parameters and distributions.

#### 5.1.4 Screening policy evaluation

For illustrative purposes, we evaluate the LYG and test burden associated with implementing population-wide screening using a hypothetical test at 5-year intervals from age 55 to 75. The test has a beta-distributed sensitivity with mean 0.9 and sample size 100 and a beta-distributed specificity with mean 0.95 and sample size 100. We sample from the parameter posterior distribution as well as the beta distribution for sensitivity to run the model and calculate a distribution of outcomes.

### 5.2 Colorectal cancer

We use DESCIPHR to simulate the natural history of CRC for a hypothetical cohort of women in the U.S. The natural history of CRC is characterized by the emergence of benign precursor lesions, or polyps, that can progress to cancer. After the development of the flexible colonoscope in the 1960s to visualize the interior of the colon and the recognition that polyp removal could interrupt the development of CRC in the 1970s [87], medical institutions began to recognize the lifesaving potential of CRC screening in the US. USPSTF guidelines recommend colonoscopy every 10 years or stool testing every 1-3 years for average-risk adults starting from age 45 as of 2021 [4]. For illustrative purposes, we assume that all cases of CRC arise from adenomatous polyps.

#### 5.2.1 Data

For background mortality, we use the annual probabilities of mortality from the 1980 female life table from the Human Mortality Database [84]. For calibration targets, we use the age-specific prevalence of adenomas from a meta-analysis of screening and autopsy studies [88], as well as the stage distribution and age-specific incidence of CRC from 1975-1979, before widespread screening for CRC in the US [85]. To simulate the time to death from cancer by stage at diagnosis, we use SEER relative survival rates from 2000 to 2021 [55]. Since stages reported for relative survival are aggregated as localized, regional, and distant, we use the relative survival of the localized stage to simulate mortality from a diagnosis at stage I, the regional stage for stages II and III, and the distant stage for stage IV.

#### 5.2.2 Natural history model

We implement a CRC natural history model with all states described in Section 2.1, including an intermediary precursor lesion state. The transitions between states are parameterized by the distributions

#### 5.2.3 Parameter estimation with BayCANN

Prior distributions for the time to lesion onset are determined using the procedure in Appendix B. For other parameters, we use a wide range of reasonable priors. Following the procedure in Appendix A.3 and multiplying the target cohort size by 1.5, we proceed with the calibration using a cohort size of 3,100,000. To generate the training sample for BayCANN, we draw 500 samples per calibrated parameter from the prior distributions using LHS for a total of 6500 parameter sets and simulate the corresponding calibration outcomes. The parameter inputs and calibration outputs other than stage distribution are scaled from 0 to 1, with 0 associated with the minimum value of each data type and 1 the maximum. 20% of the sample is held out to validate the ANN, and an 80-20 split is used for hyperparameter tuning. For training the ANN, we use a batch size of 128, a maximum number of training epochs of 5000, and a patience of 30. We perform hyperparameter tuning by randomly sampling 30% of permutations of the following hyperparameters: 1 to 4 hidden layers; 32, 64, or 128 hidden nodes per layer; including or excluding dropout layers with a dropout rate of 0.25; and reLU, tanh, or sigmoid as activation functions. The ANN architecture that minimized the loss included 2 hidden layers with the tanh activation function, 32 nodes per hidden layer, and no dropout. We run Stan with 4 chains for a maximum of 300,000 iterations and a thinning parameter of 100 to address autocorrelation, resulting in a posterior sample of 6000 parameter sets. The sample generation ran for 4.1 hours on 8 cores on a high-performance computing cluster. Hyperparameter tuning, ANN training, and Stan calibration were run on 8 cores on an Apple MacBook Pro (M1 chip, 16 GB RAM, macOS Sequoia v15.7.2), with run times of 25, 1, and 10 minutes, respectively. The calibration produced two sets of divergent chains, indicating two separate parameter regions that fit with the targets (Figure 5).

**Fig. 5:**
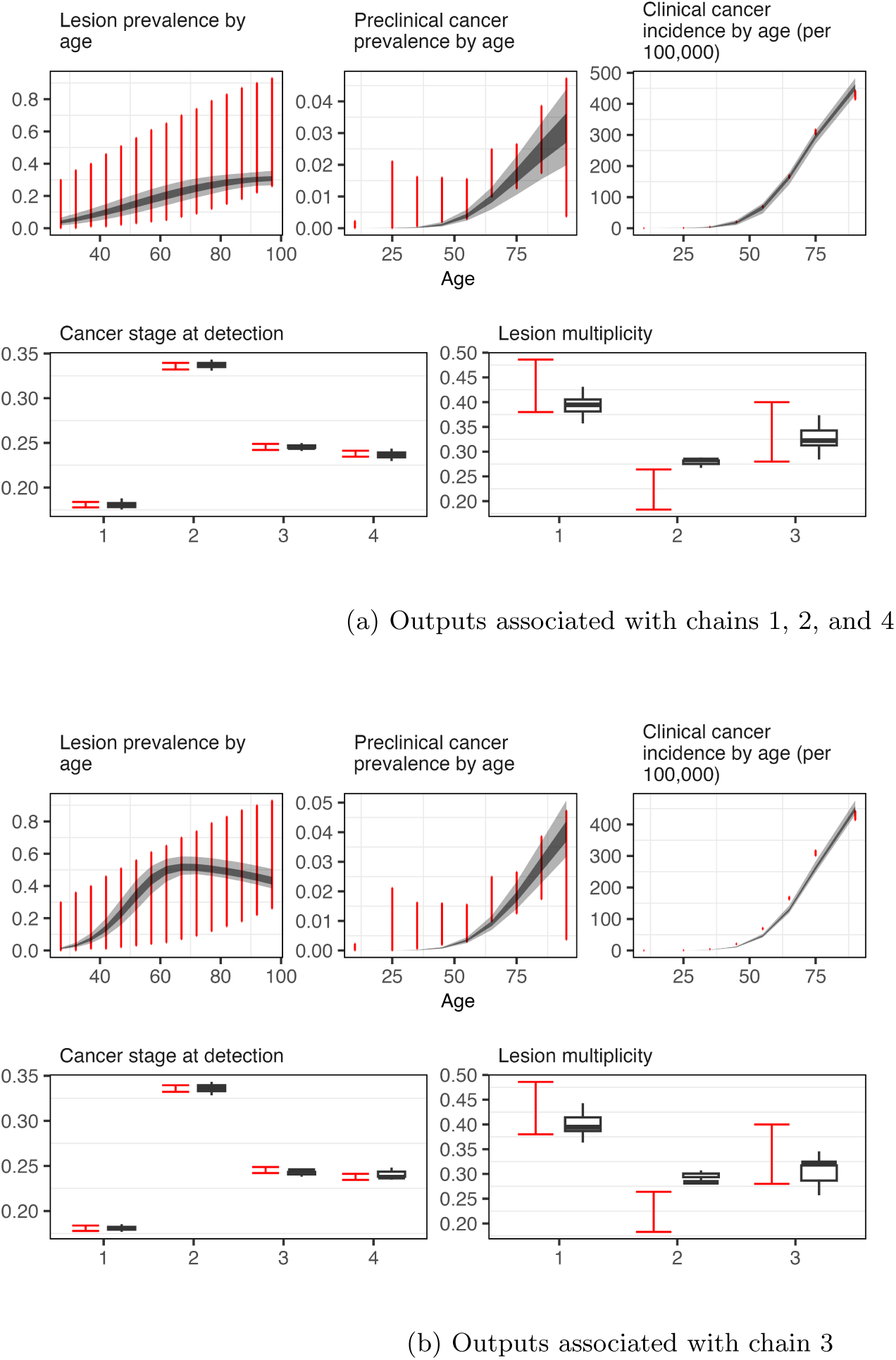
Internal validation of calibrated CRC model with model outputs in gray and 95% CIs of calibra-tion targets in red

#### 5.2.4 Screening policy evaluation

To demonstrate an application of the calibrated model, we evaluate the LYG and test burden associated with colonoscopy every 10 years from age 45 to 75. We assume a beta-distributed test sensitivity with mean 0.95 and sample size for cancer and mean 0.7 and sample size 100 for precancerous lesions. The specificity is assumed to be 100%. We sample from each region of the parameter posterior distribution as well as the beta distribution for sensitivity to run the model and calculate the distribution of outcomes associated with each region.

## 6 Discussion

We introduce the DESCIPHR modeling framework to fill a gap in open-source, flexible, end-to-end resources for implementing and deploying health decision models. The framework spans considerations for building a DES model for the natural history of cancer, estimating unobservable model parameters using Bayesian calibration, and applying the resulting posterior distributions for screening policy analysis. The DES implementation provides flexibility in modeling individual-level transitions between states compared to simpler Markov or cohort-based approaches and increases computational efficiency compared to discrete-time implementations. Improving on prior DES and calibration tutorials, the DESCIPHR codebase also demonstrates the model pre-processing and setup required to run two Bayesian calibration methods, IMABC and BayCANN, and apply the resulting joint parameter posterior distribution to make forecasts that account for the uncertainty in model parameters. We use IMABC and BayCANN because they increase efficiency relative to older Bayesian calibration techniques, such as MCMC and ABC, by leveraging adaptive sampling and metamodeling, respectively [13, 21].

In addition to linking existing modeling concepts and methods in an open-source pipeline, we make several novel methodological contributions to facilitate decision modeling with DESCIPHR. First, our target-informed approach to estimating prior distributions for the parameters governing the time to disease onset and cancer progression (described in Appendix B) circumvents the need to guess a reasonable range for a grid search manually. Despite advances in Bayesian calibration methods, assigning parameter prior distributions is still considered more of an art than a science, requiring extensive trial and error. Even though guidance suggests performing an initial grid search to find a reasonable range of parameters that could be used as priors, the grid search step still requires setting initial bounds within which to search. Initial bounds that are too narrow risk missing the target parameter space entirely. However, excessively wide bounds are prone to the curse of dimensionality; as the number of parameters and the width of the bounds increase, more samples are needed to capture a region in the parameter space close enough to the targets for calibration to be successful.

Second, our augmentations to BayCANN allow for automated hyperparameter optimization, which is a critical part of empirical machine learning because the prediction accuracy of machine learning models is sensitive to the hyperparameter configuration [89, 90]. We also add capabilities for users to distinguish output types, such as outputs that must sum to 1 versus merely range from 0 to 1, and automatically propagate their characteristics to the emulator model. These additions minimize the setup required for a modeler to train an accurate model and ensure that output constraints are satisfied. Future work could integrate more advanced and automated approaches for parameter estimation, such as reinforcement learning-based calibration [91].

To balance the learnability and adaptability of DESCIPHR, the implemented cancer natural history and screening model relies on some simplifying assumptions, only modeling a single tumor type, screening sensitivity independent of lesion size and cancer stage, and perfect adherence to screening. However, the model can be easily extended to incorporate other natural history assumptions or evaluate additional interventions based on the guidance provided in Appendix A.4. The modularity of the DESCIPHR code-base also allows researchers to easily substitute variable distributions, input data, calibration methods, and outcomes according to their needs, making the framework adaptable to specific cancer sites, as well as other diseases characterized by progressive stages that have preclinical and clinical states.

Furthermore, without targets on the stage-specific prevalence of preclinical cancer, the parameters for distributions related to cancer stage progression may not be identifiable. However, as long as the decision outcomes associated with the posteriors accurately account for the uncertainty due to nonidentifiability, a modeler may still determine the optimal decision alternative across the range of plausible parameter sets. Frameworks for decision-making under deep uncertainty can be further explored in the robust decision-making literature [92–94].

Another limitation is that comparative and external validation are not explicitly included in the DESCIPHR codebase, although the natural history and screening models can be adapted to perform them. Independently developed models have been used for comparative model validation of policy recommendations [2, 3, 95]. Unlike internal and comparative validation, external validation tests the accuracy of the model’s structure and predictions by comparing simulated outputs to empirical data not used for model construction or calibration [74–76], such as cancer mortality and incidence outcomes from screening trials [96, 97] or observational data [76, 98].

Models act as approximations of reality, and their predictive ability for decision-making carries inherent uncertainties, especially when there is insufficient data for precise calibration [8]. However, this does not belie their usefulness for predicting information that would not otherwise be measurable, especially given their history of successful inference of health trends [38, 96, 97]. For decades, simulation modeling has successfully informed cancer screening policy, though typically with bespoke, closed-source implementations [4, 95, 99]. Though this convention has the advantage of validation by consensus when diverse, independently developed models produce the same conclusion, these highly heterogeneous implementations come with a steep learning curve, a high barrier to entry, and a lack of transparency. We believe that releasing an adaptable open-source model will accelerate the progress of health decision modeling applications and methods development. As new cancer screening modalities and treatments emerge, there will be no shortage of policy decisions to make before the arrival of long-term comparative evidence. DESCIPHR represents a significant step towards streamlining model development and increasing transparency to optimally inform these decisions.

## 7 Conclusion

The DESCIPHR framework integrates the entire process of implementing a DES model for cancer natural history, calibrating the model parameters to real-world data using Bayesian methods, and applying the results to evaluate the impact of cancer screening strategies. The modular organization of the DES model and pipeline, the model’s flexibility in accepting parametric and non-parametric inputs, and the demonstration of the pipeline with a ground truth simulation facilitate downstream users’ ability to understand the pipeline and customize it to their modeling specifications. We provide and illustrate all steps of DESCIPHR in an open-source code repository as a foundation for developing, calibrating, testing, and deploying future decision models for cancer and other progressive diseases.

## 8 Declarations

### 8.3 Availability of data and material

All data generated for this manuscript are provided in a GitHub repository accessible at https://github.com/sjpi22/tutorial cancer modeling des.

### 8.4 Ethics approval

Not applicable.

### 8.5 Consent to participate

Not applicable.

### 8.6 Code availability

All code used for this manuscript is provided in a GitHub repository accessible at https://github.com/ sjpi22/tutorial cancer modeling des.

### 8.7 Author contributions

Conceptualization, methodology: FAE, CMR, SP, JGF. Data curation, validation, writing — original draft: SP. Formal analysis: SP, FAE, CMR, CPA. Project administration: FAE, SP, CMR. Resources: JGF, FAE, JHC. Software: SP, CPA, FAE, CMR. Supervision: FAE, CMR, JHC, JGF. Visualization: SP, CPA, FAE. Writing — review and editing: all authors.

## Data Availability

All data produced are available online at

https://github.com/sjpi22/tutorial_cancer_modeling_des

## Acknowledgments

We are grateful to Michael C. Higgins, PhD, for feedback on the natural history model. Additionally, some of the computing for this project was performed on the Sherlock cluster. We would like to thank Stanford University and the Stanford Research Computing Center for providing computational resources and support that contributed to these research results.

## A. Adapting the model

### A.1 Model configurations

The YAML files in the configs folder provide a central location for model structure specifica-tions. Under params model, parameters that serve as inputs to the load model params() function in R/01 model inputs functions.R can be defined, such as the indicator for whether to include a pre-cancerous lesion state (lesion state) and the list of cancer stages (v cancer). Similarly, customized parameters for IMABC, BayCANN, Monte Carlo error analyses, target coverage analyses, and screening strategy analyses can be set respectively in params imabc, params baycann, params montecarlo, params coverage, and params screening.

Each random variable in the simulation model may be customized by providing the distribution name, parameters, and source in a list. For example, we may create a list object named d time H L for the distribution of the time from the healthy state to the first precancerous lesion as follows, using the base R accelerated failure time parametrization of the Weibull distribution:

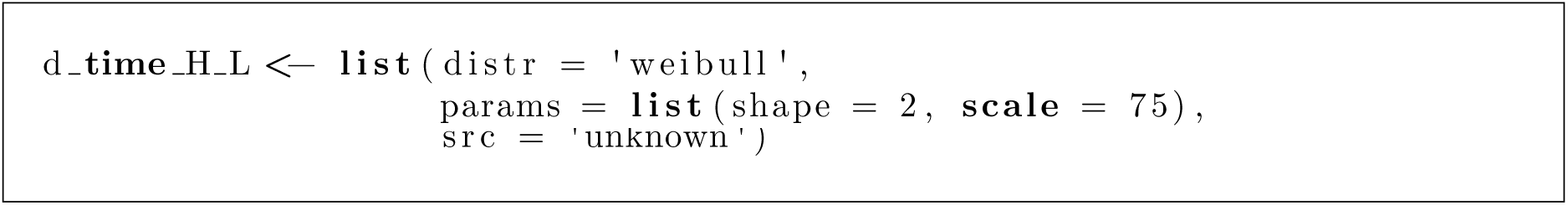

With this framework, the user may substitute different parametric or empirical distributions for each of the TTE variables. We may then draw 10 samples from the distribution using the query distr function as follows:

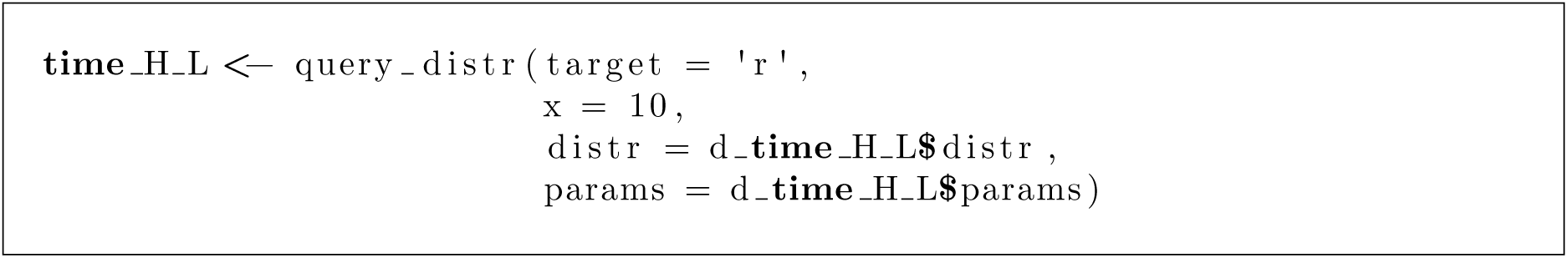

The query distr() function appends the target letter to the distribution name and calls the result-ing function with x as the first input and params as the following inputs, following R’s syntax for probability distributions, in which d indicates the density, p the probability, q the quantile, and r random generation. As a result, the value of distr should be a recognized function when appended to one of the four values of target; acceptable values corresponding to R base distributions include exp, gamma, and norm, while some functions may be loaded from other packages (for instance, functions for the Gompertz distribution are available from the VGAM or flexsurv packages).

We have also provided functions to sample from empirical distributions in the file R/utils/distr empirical.R. The required input data include an ordered discrete variable xs, such as an age interval, and the corresponding probability mass probs. The function dempirical() retrieves the value of the probability mass function for the interval containing the queried value. The function pempirical() calculates the cumulative probability by summing the values of probs up to the queried value of xs. Its inverse is qempirical(), which outputs the corresponding quantile of a probability input. The random generator function rempirical() samples from the categorical variables weighted by the probability mass. For continuous variables such as age, the function includes an option to apply a uniform correction to achieve values in between the discrete values of xs and probs.

To modify the time-to-event distributions used in the simulation model, the user may do the following:

1. Specify distributions in a CSV file such as data/priors.csv, which updates the distributions of the natural history model accordingly when used as an input to load calib params() in 03 calibration general functions.R as file priors or to load calib params() in R/01 model inputs functions.R as file.distr
2. Manually change the distributions after loading the model parameters.
3. Change the default distributions set in 01 model inputs functions.R/load model params()

### A.2 Data

To run an analysis with custom input data and calibration targets, replace the default datasets in the data folder with custom data and make the following modifications according to the type of input.

1. Model inputs (background mortality, relative survival)

a. Update the data file paths for background mortality (file.mort) and relative survival (file.mort) in one of the YAML files in the configs folder under params model.
b. If necessary, modify load lifetables() for the background mortality data and load surv data() in R/utils/data processing.R for the relative survival data so that the data inputs can respectively be converted to probability distribution objects using set mort distr() and set surv distr(), also defined in R/utils/data processing.R. These functions are all called in load model params(), which is defined in R/01 model inputs functions.R and loads the cancer natural history model parameters.
2. Calibration targets

a. Replace or add parameters for calibration targets under params calib:l params outcome for non-lesion calibration targets, such as prevalence of preclinical cancer, incidence, and stage distri-bution, and params calib:lesion state true:l params outcome for precancerous lesion-specific calibration targets in the the applicable YAML file in the configs folder. Each calibration target should be associated with a uniquely labeled list of parameters, including

i. file path: path to target data
ii. outcome type: label for target type, which indicates the functions to load the data in R/utils/data processing.R (load [outcome type]()) and calculate the analogous outcome from simulated data in R/utils/epi functions.R (calc [outcome type]())
iii. categorical: indicator for whether the target is a categorical distribution (i.e., percentages that must sum to 1)
iv. get params: named list of inputs to calc [outcome type]() that must be retrieved using get()
v. lit params: named list of literal inputs to calc [outcome type]()
b. If necessary, modify or create functions of the form load [outcome type]() in R/utils/data processing.R to load and process each type of target. The output should include the following columns: “target names” with unique labels for each target, “target groups” to label the entire category of outcomes, “target index” for the indices associated with each value of the target, “targets” for the target value at each index, and “se” for the standard error
c. For any new types of calibration targets, create functions of the form calc [outcome type]() in R/utils/epi functions.R to calculate the corresponding targets from a matrix of simulated patient trajectories

### A.3 Choice of cohort size

When implementing an individual-level simulation model, a key question is how many life trajectories to simulate. Selecting the cohort size involves similar considerations as choosing a sample size in prospective trials; as the cohort size increases, the Monte Carlo error of the summary outcome estimates decreases [31], but the time required to run the simulation increases [29]. The acceptable Monte Carlo error of the summary statistics generated depends on the task at hand (e.g., calibration, validation, policy evaluation). To facilitate convergence during calibration, the Monte Carlo error of the simulated outcomes should not exceed the estimation error of the corresponding calibration targets [29]. For strategy evaluation, the Monte Carlo variability should be much smaller than the mean differences in outcomes between strategies [100].

To determine the minimum sample size *N_target_* required for Monte Carlo variability to be lower than each target’s standard error *SE_target_*, a modeler could first calculate the minimum standard error *SE_n_* of the outcomes of interest with a small preliminary sample *n*, such as *n* = 1, 000, then solve *N_target_* = *n*(*SE_n_/SE_target_*)^2^, as the standard error and the square root of the sample size are inversely proportional [100]. The modeler may then analyze the computation time for varying sample sizes to determine a cohort size reasonably larger than *N_target_* that falls within their computational limits [29]. One study alternatively determined its microsimulation cohort size by performing 20 simulations each for sample sizes starting at 100,000 and incrementing by 100,000 until the standard deviation of the parameter estimates no longer decreased meaningfully, thus achieving near-minimum Monte Carlo uncertainty [101].

### A.4 Further augmentations

This section describes ways to adapt the current model implementation to accommodate additional cancer natural history assumptions and interventions. For an overview of where to add functions and parameters and how to specify model configurations to support these adaptations, refer to Appendix A.2.

#### A.4.1 Additional states

Modeling certain cancer types or evaluating certain non-screening interventions may require additional states from those that are currently implemented in the model. For example, infection may precede precancerous dysplasia or cancer onset in the case of human papillomavirus for cervical cancer [56], hepatitis B or C for liver cancer [102], and *H. pylori* for gastric cancer [103]. Thus, modeling interventions to prevent or treat such infections may require adding a state to reflect reduced transition rates to cancer through that pathway. Furthermore, to evaluate surveillance strategies for cancer recurrence or therapies based on the line of treatment, states for recurrence and detection of recurrence must be added between initial diagnosis and death from cancer.

Additional states in the natural history process can be integrated into the code by performing the following steps:

1. Updates to 01 load model inputs.R

a. Assign a label for the new state as a letter different from existing state labels (H, L, P, C, and D) and assign its order relative to other states.
b. Add a binary indicator to the arguments of the load model params() function for the inclusion of the state in the decision model, analogous to the indicator for the precancerous lesion state (L).
c. Update the vector of states v states to include the new state if the indicator argument for the state is TRUE.
d. Create placeholder lists for relevant probability distributions for time-to-event variables leading to and from the new state and state. Wrap the lists in an if-statement that checks whether the state label is in v states. Adjust the code to default to the correct placeholders if the state is not included.
2. Updates to 02 decision model functions.R

a. Create one or more subfunctions to generate the events and characteristics related to the new state, including variables that lead into the new state and the following state (for example, one could create a simulate infection onset() function to simulate the time of a precipitating infection). The function(s) should take the patient matrix m times and model parameters l params all as inputs. If the new state progresses into the lesion or preclinical cancer state, update simulate disease onset() to integrate the time of disease onset with the state.
b. Call the subfunctions in the main function run base model() in the desired order of the states. Wrap the subfunctions in an if statement calling them if the new state is in v states.
3. Sanity checks

a. Load model parameters and check that the distributions for time-to-event variables connecting to and from the new state exist.
b. Run the model with the loaded parameters and check that the patient-level matrix outputs contain the time-to-event variables. Check that they sum up as you would expect.

#### A.4.2 Risk factors

Risk factors that are treated as time-invariant in the model, such as insurance status, family history, or geographic region at the time of screening initiation, may be added in one line to the simulate baseline data() module. Their population distributions, as well as their interactions with other distributions’ parameters (for example, as covariates in the hazard of cancer onset) may be specified in load model params().

For time-varying risk factors, such as smoking behaviors [104], a module can be added using the steps in A.4.1 to simulate the trajectory of the risk factor over each individual’s lifespan. A separate module can be added to specify how disease risk or mortality changes with the risk factor, as implemented by the CISNET Lung Group models [95].

#### A.4.3 Additional subtypes

Heterogeneity in precancerous lesion pathways, as in colorectal cancer [105], or cancer subtypes, such as for breast cancer [38], may be important to model if associated with differential outcomes. To simulate multiple precancerous lesion types, one may sample an onset time and lifetime lesion count for each type in the simulate disease onset() subfunction, potentially with an individual lesion risk index to induce correlation. Then, in the simulate additional lesions() subfunction, one may loop over the lesion types to simulate the time to cancer progression using type-specific progression rates. In the screening module, the screening sensitivity inputs and code for sampling positive test results can be modified to use different detection probabilities by lesion type.

Assuming that individuals only develop one of several mutually exclusive cancer subtypes in the model, one may sample from the observed distribution of subtypes from cancer incidence data, then sample an age of onset by subtype. Differential cancer progression rates, symptomatic detection rates, survival, and intervention efficacy may also be specified based on subtype. If multiple subtypes of cancer per individual are modeled, one may generate event times starting from cancer onset at the tumor level, as is done for precancerous lesions.

#### A.4.4 Lesion size, stages, and regression

Precancerous lesion or tumor size may be important to model if screening test sensitivity varies with lesion size, as in the case of stool tests for colorectal cancer [106]. The trajectory of lesion growth can be modeled using discrete size categories with specified transition distributions between categories. Similar methods can be used to model precancerous lesion stages, such as cervical intraepithelial neoplasia (CIN) 1, 2, and 3 [107]. Tumor growth has also been modeled in continuous time using exponential, Gompertz, and Janoschek curves [12, 28, 50]. Parameters for transition distributions and growth curves can be calibrated to real-world targets on the distribution of lesion size. In the screening module, a function for the test sensitivity based on lesion size would be added as an input so that the probability of detection based on the lesion size at the time of the screening test can be set.

In addition, some cancer types, such as cervical cancer [107, 108], show evidence of precancerous lesion regression. To model regression, after instantiating lesions, one can sample a binary variable indicating whether a lesion will regress or progress. If modeling lesion size categories, one may sample the times to the maximum lesion size and transitions to smaller categories until the lesions that will regress no longer exist. Continuous trajectories for lesion shrinkage over time can also be used. In the screening module, one can either assign a test sensitivity of zero for lesions that are too small to be detected or exclude lesions that have completely regressed from screening.

#### A.4.5 Incomplete resection

To model incomplete resection of precancerous lesions during an invasive screening or diagnostic test, the probability of incomplete resection, the distribution of the size of the remaining lesion, the growth trajectory after resection, and assumptions for cancer progression after resection would need to be added as inputs or otherwise specified. Using these parameters, for lesions detected at each screening, one could sample whether the lesion is fully resected, flag completely resected lesions as removed, reset the size of incompletely resected lesions, sample a new time to preclinical cancer for incompletely resected lesions, and calculate the lesion size at the next screening test.

#### A.4.6 Adherence

Given the imperfect real-world adherence to U.S. cancer screening guidelines [109–112] and differential preferences in screening modalities [113, 114], there is increasing interest in adherence as a parameter in models evaluating screening strategies, such as in [115]. In the simplest case, adherence may be modeled as a single probability that is applied to each individual and independent between tests, though this may not be realistic. To implement this form of adherence, at each time that individuals are due for screening, before sampling test results, one may sample whether individuals participate and proceed with calculating test outcomes (e.g., false positives, detection of lesions or cancer) only among adherent individuals. Alternatively, at the time that individuals are due for screening, one may sample from a time-to-event (TTE) distribution for the delay in testing to simulate realistic uptake patterns. To incorporate heterogeneity in screening adherence, one can assign individuals to categories of adherence and either set a fixed screening interval or sample from separate TTE distributions for each category. For example, in [116], individuals are categorized as annual screeners, biennial screeners, and irregular screeners, each with separate survival curves for the time to next breast cancer exam.

#### A.4.7 Additional decision outcomes

Though screening test costs are incorporated in the pipeline already, more outcomes, such as quality-adjusted life years (QALYs) and cancer treatment costs, may be needed for a comprehensive cost-effectiveness analysis, weighing the financial burden of screening programs against the life extension and cost savings of screening-mediated cancer prevention or early detection. Since the DES model already simulates the time in each health state, one can apply state-specific utility weights to calculate QALYs as well as annual costs of living in a particular health state. At the time of each screening test, one can also sample the occurrence of screening-related adverse events and their associated costs.

## B. Target-informed priors

For an acyclic disease model with progressive states, we show how to quantitatively derive prior distributions for the parameters governing the time to disease onset given epidemiological data for each state. In the cancer setting, we take data on the incidence of clinical cancer *I_C_*(*t*) and prevalence of preclinical cancer *P_P_* (*t*) across multiple age ranges. The midpoint of the *i*-th age range is denoted by *t_i_*. First, we derive the CDF of clinical cancer *F_C_*(*t*) from the incidence rates. Next, we use the CDF of clinical cancer and the prevalence of preclinical cancer to estimate the CDF for the time to preclinical cancer onset *F_P_* (*t*). The same methods can be used to derive the CDF for precancerous lesion onset *F_L_*(*t*) given data on the prevalence *P_L_*(*t*) of precancerous lesions by age. In this appendix, we also discuss how cancer stage distribution targets can inform the parameter priors for cancer stage progression.

### B.1 Deriving the CDF of clinical cancer from the incidence rate

The National Cancer Institute (NCI)’s Surveillance, Epidemiology, and End Results (SEER) Program calculates cancer incidence as the number of new cases in a year divided by the total population [117]. Using the cobs R package, we fit a constrained B-spline *Î_C_* (*t*) to the incidence values *I_C_*(*t_i_*) at each midpoint of the age range *t_i_*, with knots at 0, *t*_2_, every 3rd midpoint after *t*_2_, and the maximum upper bound of the age ranges. The knots are sparse to prevent overfitting. For improved extrapolation beyond the data ranges, constraints can be set so that the fitted spline is negative at 0 (to prevent high incidence rates at 0), increasing, or nonnegative. Since incidence is nonnegative, any negative values of the fitted spline are assumed to be set to 0 in *Î_C_* (*t*).

To convert the SEER incidence to a hazard rate or probability density that can be integrated to calculate the CDF, we must respectively subtract the living clinical cancer cases from the denominator or add the deceased clinical cancer cases to the denominator. Both methods would require calculating death rates from cancer, so we proceed with the latter, which is more direct. Letting 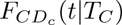 denote the CDF for the time from cancer diagnosis to death from cancer conditional on the age at diagnosis, we estimate the probability 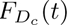 of dying from cancer by age *t* by integrating over possible diagnosis and death times using

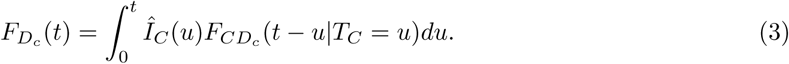

We then calculate the probability density of clinical cancer *f_C_*(*t*) by scaling the clinical cancer incidence rates by the proportion of people who have not died from clinical cancer using

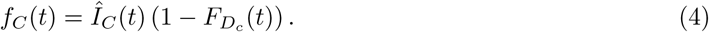

Finally, the CDF is calculated by integrating over the PDF using

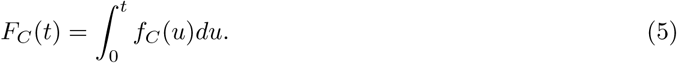

The risk of death from other causes is not considered because we assume that it is independent of the cancer disease process.

We can derive a crude estimate of 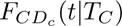 for deriving priors assuming that relative survival rates are independent of age at diagnosis and are exponentially distributed. To calculate the rate of cancer death, we start with SEER data on relative survival over 10 years by stage *s* at diagnosis, denoted as 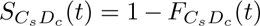. We extrapolate relative survival beyond 10 years by fitting constrained splines or parametric distributions to produce estimates 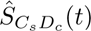. The average relative survival for each stage, 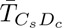 is estimated using a Monte Carlo simulation from 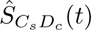, and the overall average time to death from cancer 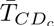 weights the stage-specific averages by the proportion of the population diagnosed at each stage 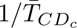.serves as the exponential rate for *F_CDc_* (*t*).

Alternatively, if the incidence rate excludes prior clinical cancer cases from the denominator, it is equivalent to the hazard rate of clinical cancer [118]. The fitted spline for the incidence rate can then be integrated directly to calculate the cumulative hazard *H_C_*(*t*) and the CDF as follows:

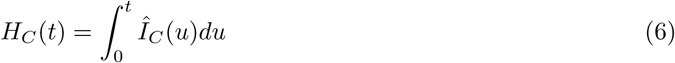

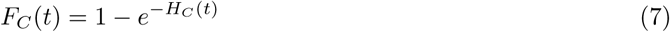

### B.2 Deriving the CDF of disease onset

Prevalence data are assumed to come from screening studies that exclude patients already diagnosed with cancer. To calculate the cumulative probability of preclinical cancer, we must therefore rescale preclinical cancer prevalence as a proportion of the whole population and add the proportion of people who have passed the preclinical cancer stage as follows:

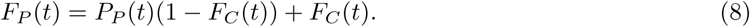

Using the 95% CIs of incidence and prevalence, we calculate lower and upper bounds for the CDF of the time to cancer onset.

If precancerous lesion prevalence is similarly derived from screening studies that include preclinical cancer but exclude clinical cancer cases, we can likewise calculate the CDF for precancerous lesion onset using:

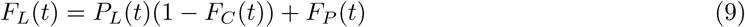

Figure 6 demonstrates the results of this algorithm, showing that the ground truth simulated CDFs (solid lines) for precancerous lesion (green), preclinical cancer (orange), and clinical cancer (red) onset are within the 95% CIs (dashed lines) of the respective CDFs (points) estimated from the prevalence and incidence targets.

Denoting *F_X_* (*t*) as the CDF estimate for the time from birth to disease onset, whether it be pre-cancerous lesion or preclinical cancer onset, we let 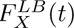 and 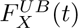 be the lower and upper bounds, respectively. Using these CDFs, we may fit parametric distributions and derive bounds for a plausible range of parameters. In our example, we assume that *F_X_* (*t*) follows a Weibull distribution. Since the CDF of a Weibull distribution equals 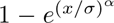, where *α* and *σ* are the shape and scale parameters, respectively, we can perform a weighted least squares regression by transforming the CDF equation into

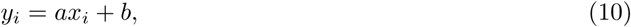

where *y_i_* = ln(− ln(1 − *F_X_* (*t_i_*))), *a* = *α*, *x_i_* = ln *t_i_*, and *b* = −*α* ln *σ* [119]. Each point is weighted by 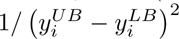, where 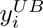 is the transformation *y*_i_ applied to 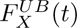 and likewise for 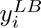. This weighting scheme downweights values of the CDF for which there is less certainty as reflected by wider CIs. With the coefficient *a* and intercept *b* from the regression, we solve for *α* = *a* and *σ* = *e*^−^*^b/a^*. Heteroskedasticity-robust standard errors of the regression parameters are used to derive 95% CIs of the parameters, which are converted to bounds for *α* and *σ*. The final bounds of the priors for *α* and *σ* are calculated by expanding the CI bounds by 20% to ensure that the targets would be covered. As shown in Figure 7, the range of priors for the parameters governing the time to disease onset contains the ground truth parameters.

**Fig. 6:**
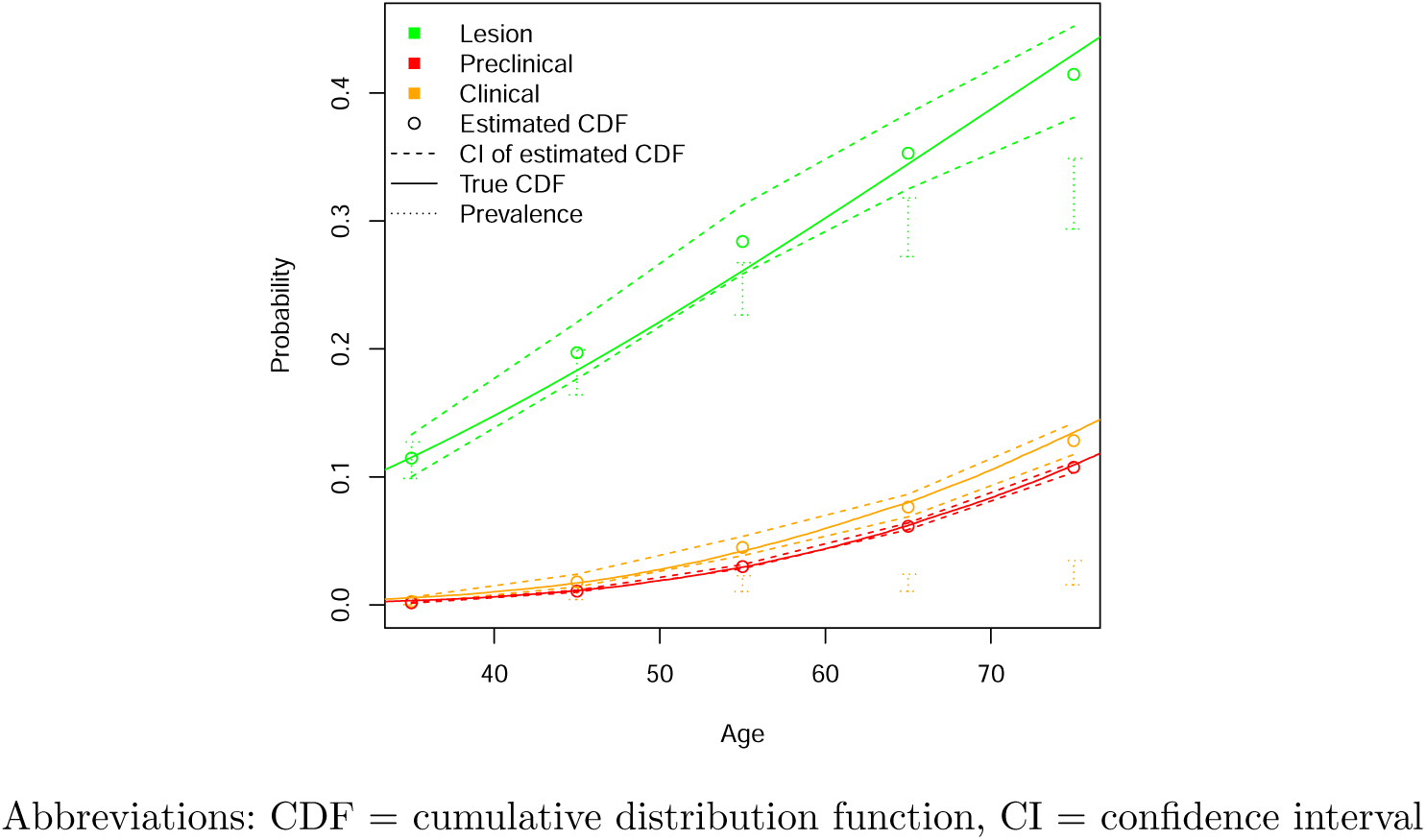
Coverage of ground truth simulated CDFs by the CDF estimates

**Fig. 7:**
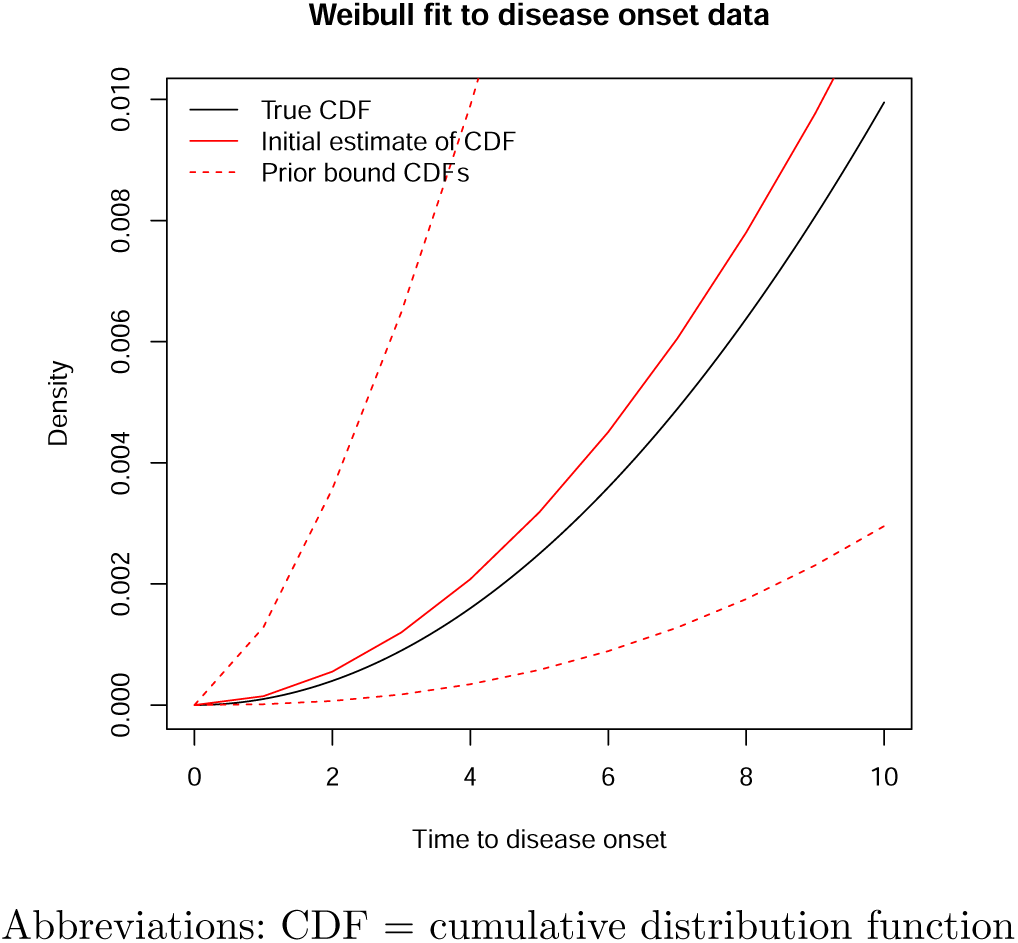
Coverage of ground truth Weibull distribution by priors

### B.3 Deriving priors for cancer progression

Rather than setting independent priors for the rates 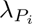 of the preclinical cancer stage progression TTE variables 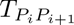, we make two alterations to the model parametrization for calibration to take advantage of intuition about cancer biology and model structure. First, we set a prior for 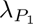 and define new parameters *h_i_* as the unknown hazard ratios (HRs) applied to 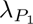 to calculate 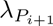. For example, HR priors that are set to range from 0.7 to 2 indicate a belief that the average progression time for each stage is no more than twice as fast as that of the previous stage and no less than 30% slower.

Next, we leverage that for independent exponential variables 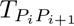 and 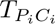 with respective rates 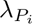 and 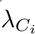,

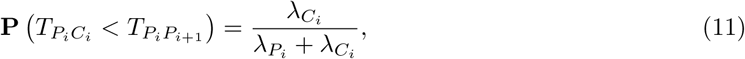

where 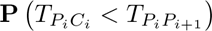 is equivalent to the proportion of cancers detected at stage *i* conditional on having reached preclinical stage *i*. Given a true stage distribution 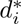 that would be observed in the absence of death from other causes,

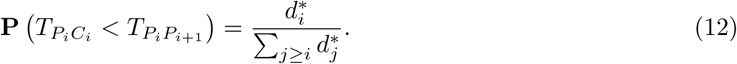

However, we expect the observed stage distribution *d_i_*to differ from 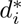 because later stages are more likely to occur at older ages and are more likely to be censored, resulting in a slight overrepresentation of earlier stages in the observed distribution. We thus define unknown parameters 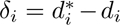 to correct for this difference, solving for 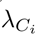 in Equation 11 by substituting the left-hand side using Equation 12 and 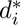 for *d_i_* + *δ_i_*. The constraint that the stage distribution must sum to one removes a degree of freedom, so *δ_i_*is fixed at 0 for the final stage. Otherwise, prior distributions for *δ_i_* are set at small values, such as between −0.05 and 0.05, with earlier stages weighted towards more negative values.

## C. Testbed model

To illustrate the effectiveness of the calibration approaches, we simulate calibration targets using a ground truth cancer model and assess whether the two Bayesian calibration methods recover the ground truth parameters when calibrating to the simulated targets.

### C.1 Data

We simulate data using a ground truth cancer natural history model that includes a precancerous lesion state and 4 stages of cancer, assumes a 50% probability of being male, and follows the variable distributions in Table 3. We first run the ground truth model to generate targets for evaluating the calibration process, including precancerous lesion prevalence, preclinical cancer prevalence, clinical cancer incidence, and distributions of lesion multiplicity and cancer stage at diagnosis. The prevalence targets are summarized for 10-year intervals from age 30 to 80, lesion multiplicity from age 50 to 80, and incidence for 10-year intervals from age 30 to 90. All individuals diagnosed with cancer in their lifetimes inform the cancer stage distribution. For the prevalence and multiplicity targets, we sample an observation date at which the presence of disease or number of lesions is assessed for each individual uniformly within the age ranges, and individuals censored due to cancer diagnosis or death before the observation age are excluded. For the multiplicity targets, we consider the percentage of screened individuals with 1, 2, or 3+ lesions out of those with any lesions. In total, there are 23 targets. The ground truth simulation also outputs relative cancer survival for the first ten years after diagnosis stratified by the stage at diagnosis.

**Table 3:** Testbed ground truth model parameters.

| Variable | Description | Distribution | Value |
| --- | --- | --- | --- |
| $M$ | Male sex | Bernoulli | 0.5 |
| $T_{H,D_o}$ | Time from birth to death from other causes | Gompertz<br>(CDF: $1 - e^{-a/b(e^{bt}-1)}$) | $a_{male} = 0.000078$,<br>$b_{male} = 0.08$,<br>$a_{female} = 0.000076$,<br>$b_{female} = 0.078$ |
| $T_{C_i,D_c}$ | Time from stage $i$ clinical cancer to death from cancer | Exponential<br>(CDF: $1 - e^{-r_i t}$) | $r_1 = 0.05$, $r_2 = 0.08$, $r_3 = 0.16$, $r_4 = 0.5$ |

**Table 3:** Testbed ground truth model parameters
| Variable | Description | Distribution | Ground truth value |
| --- | --- | --- | --- |
| $T_{H,L}$ | Time from birth to onset of first lesion | Weibull<br>(CDF: $1 - e^{-(t/\sigma)^\alpha}$) | $\alpha = 2$, $\sigma = 100$ |
| $N_L$ | Number of additional lesions before death from other causes | Poisson with rate $\nu$ for time to next lesion | $\nu = 0.0625$ |
| $T_{L,L_j}$ | Time from first lesion to onset of additional lesion $j$ | Uniform from 0 to $T_{H,D_o} - T_{H,L}$ | N/A |
| $T_{L_j,P}$ | Time from onset of lesion $j$ to conversion to preclinical cancer | Weibull<br>(CDF: $1 - e^{-(t/\mu)^\kappa}$) | $\kappa = 3$, $\mu = 50$ |
| $T_{P_i,P_{i+1}}$ | Time from stage $i$ to $i+1$ of preclinical cancer | Exponential with rate $\lambda_i$ | $\lambda_1 = 0.33$, $\lambda_2 = 0.4$,<br>$\lambda_3 = 0.6$ |
| $T_{P_i,C_i}$ | Time from stage $i$ preclinical cancer to stage $i$ clinical cancer | Exponential with rate $\gamma_i$ | $\gamma_1 = 0.09$, $\gamma_2 = 0.02$,<br>$\gamma_3 = 0.5$, $\gamma_4 = 0.5$ |

### C.2 Natural history model

For calibration and decision analysis, we use a natural history model with the same model structure, distributional forms, and background mortality distribution as the ground truth model. The model includes 13 parameters for 11 probability distributions that must be estimated via calibration. To determine the cohort size for calibration, we use an initial sample of 50,000 and conduct 50 simulations to estimate the Monte Carlo error of the prevalence, incidence, and stage distribution outputs. The required sample size for the standard deviation of the outputs to be less than the standard errors of all targets is doubled and rounded up, resulting in a cohort size of 300,000.

### C.3 Parameter estimation with IMABC and BayCANN

Uniform prior distributions for the calibrated parameters are created by randomly sampling a lower bound between 16% and 80% of the ground truth value and an upper bound between 120% and 216% of the ground truth value. This procedure generates wide bounds representative of what might realistically be used for Bayesian calibration. The priors for the parameters for time to lesion onset are refined using the procedure described in Appendix B.

We calibrate the model separately with IMABC and BayCANN. For the IMABC parameters, we use the (1 − 10^−15^) × 100% CI of the calibration targets for the starting bounds, the 95% CI for the stopping bounds, 1000 initial samples per unknown parameter, 10 normal mixtures, 50 points per normal mixture, and a target posterior ESS of 1000. For our example, the posterior distribution ultimately contains a total of 1067 parameter sets. For BayCANN, we use the same ANN training scheme and hyperparameters described in the colorectal cancer model example in Section 5.2.3. In Figure 8, the box plots of simulated

**Fig. 8:**
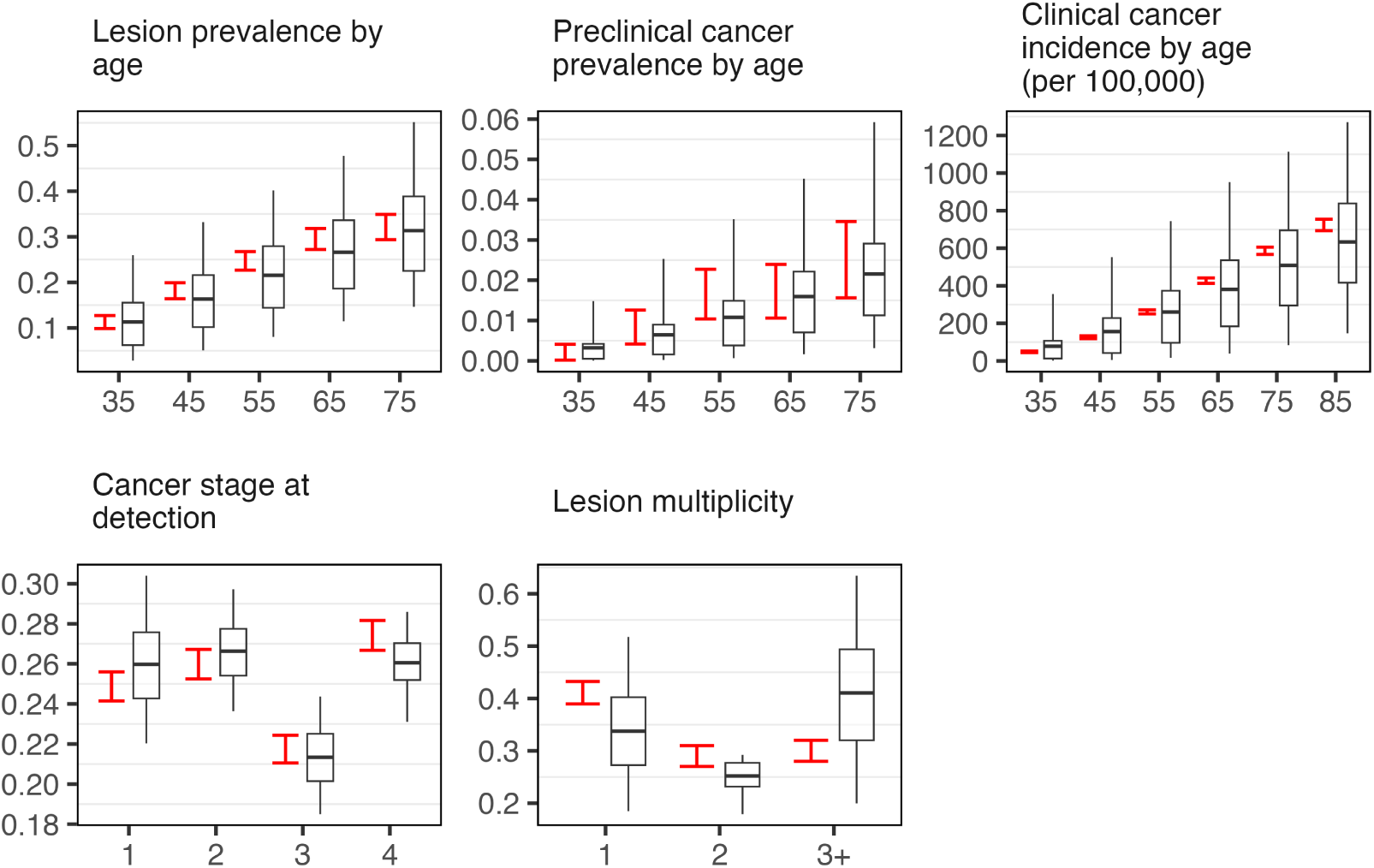
Target coverage of prior distributions, with box plots corresponding to simulated outputs and 95% CIs of targets in red outputs associated with a LHS of the parameter prior distributions cover most of the corresponding target 95% CIs in red.

Figure 9 demonstrates the ability of the IMABC and BayCANN posterior distributions to produce model outputs in gray consistent with the calibration targets in red. Although ground truth parameter values are unavailable in practice, we further demonstrate the accuracy of the calibration algorithms for this example by showing the overlap of the posterior distribution histograms and the true parameters, represented by the red vertical lines, in Figure 10.

**Fig. 9:**
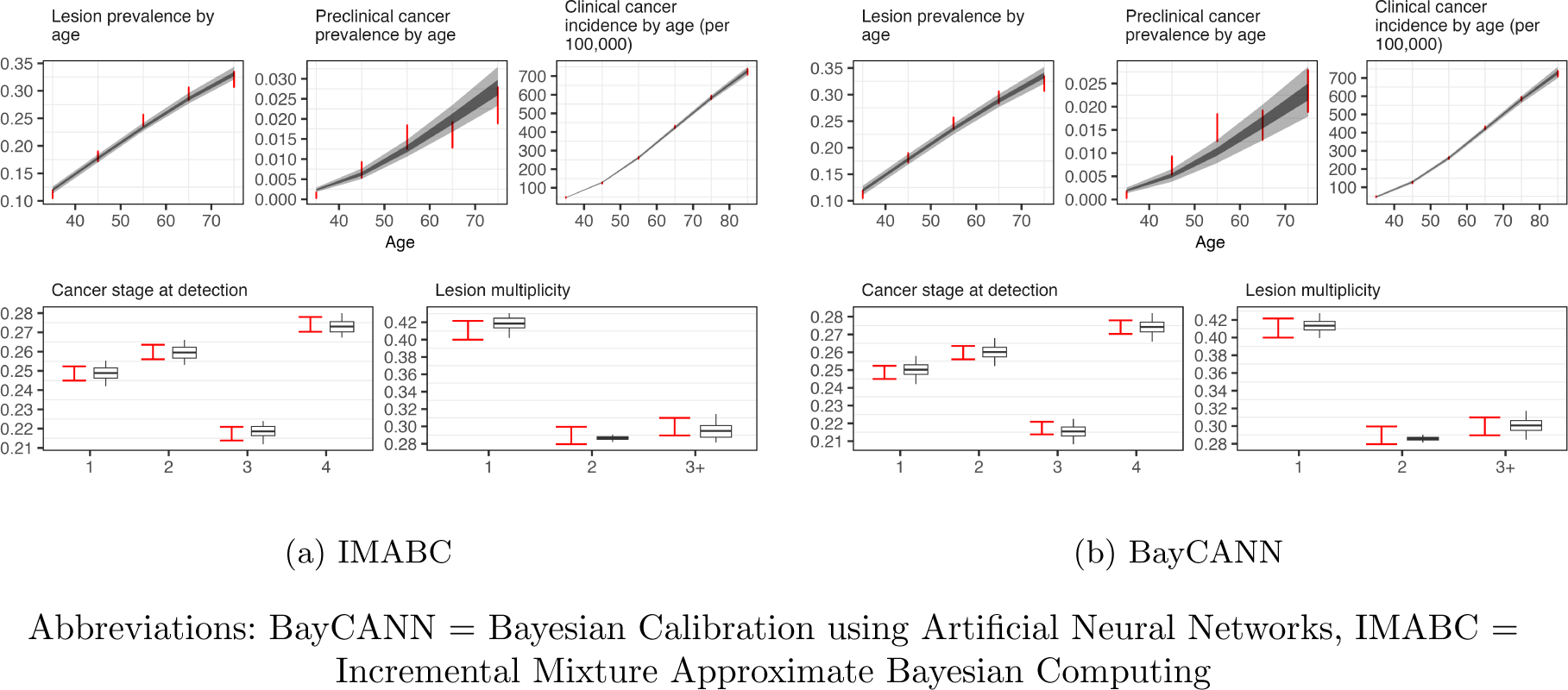
Internal validation of posteriors with model outputs in gray and 95% CIs of calibration targets in red

**Fig. 10:**
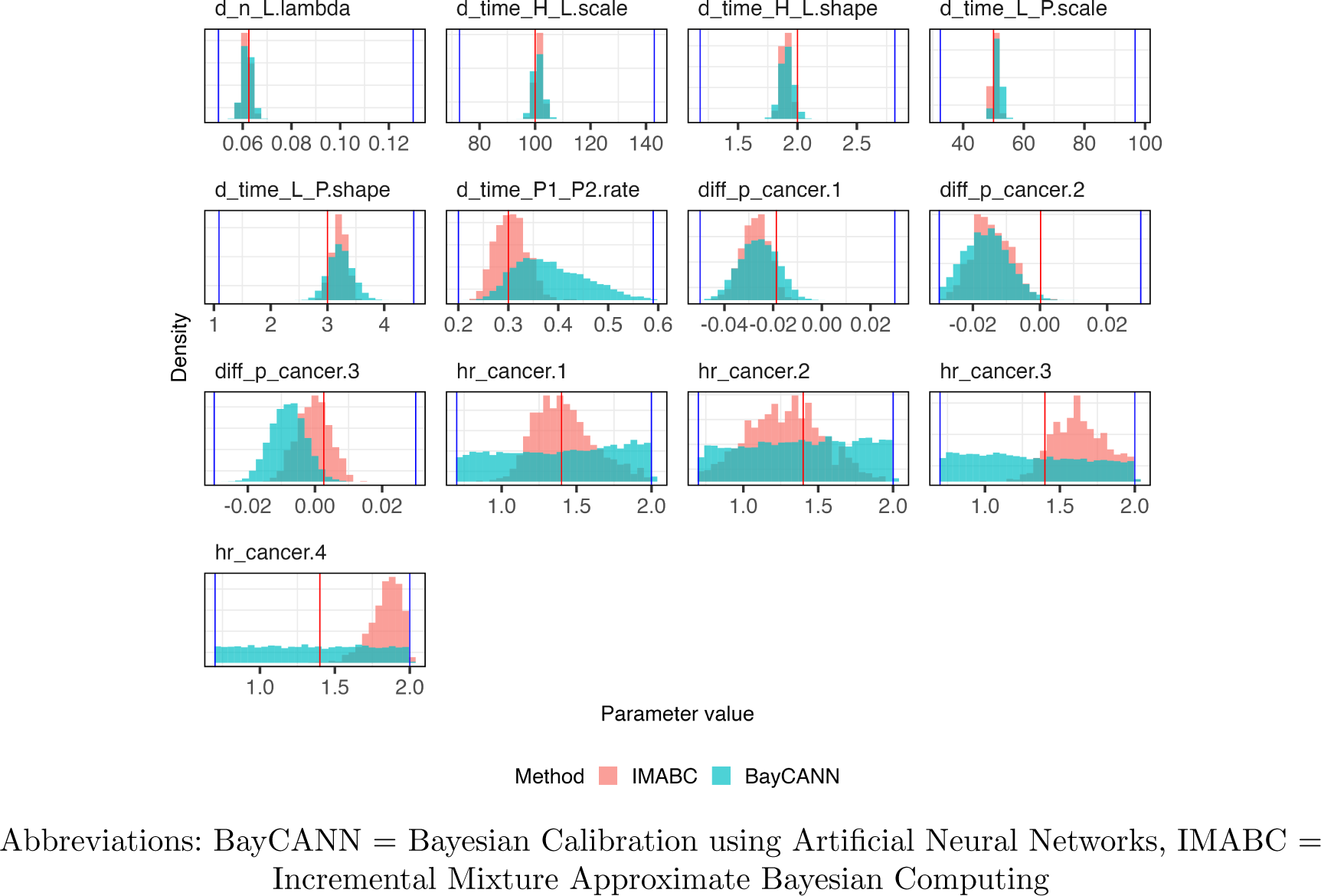
Posteriors (blue and red histograms) versus prior bounds (blue vertical lines) and true parameters (red vertical line)

The unimodal likelihood profiles for parameters such as the shape and scale of the Weibull distribution for the time to lesion onset indicate that the calibration algorithms converged to a unique solution [77]. However, the wide range and high correlation (see Figures 11 and 12) of other parameters, such as the hazard ratios for the rates of preclinical cancer progression, indicate that not all parameters are identifiable given the available targets. This is because the preclinical cancer prevalence and clinical cancer incidence targets inform the time from cancer onset to detection but are not fine-grained enough to inform the time between stages of preclinical cancer. These targets are consistent with both faster progression rates in earlier stages combined with slower progression in later stages and vice versa, resulting in non-identifiability.

**Fig. 11:**
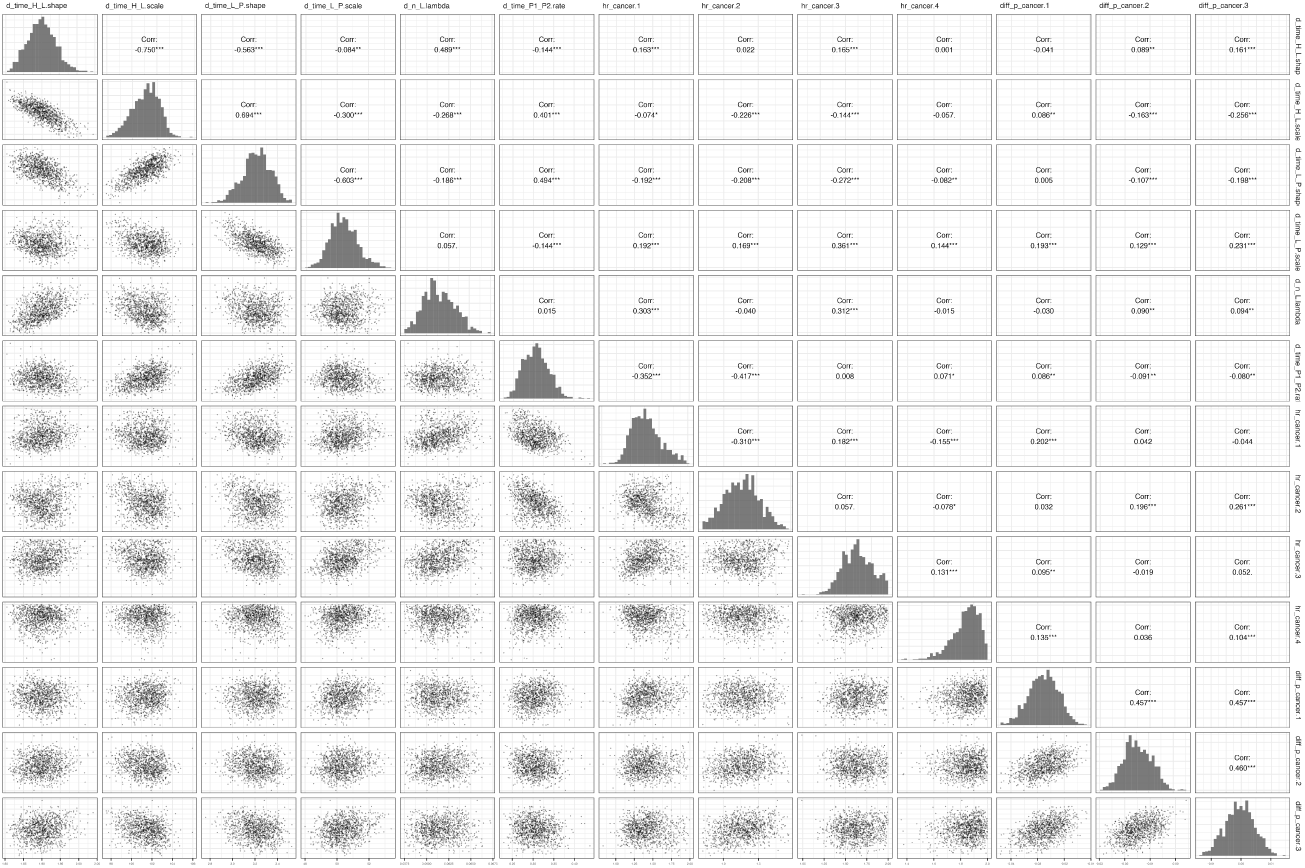
IMABC correlation plot

**Fig. 12:**
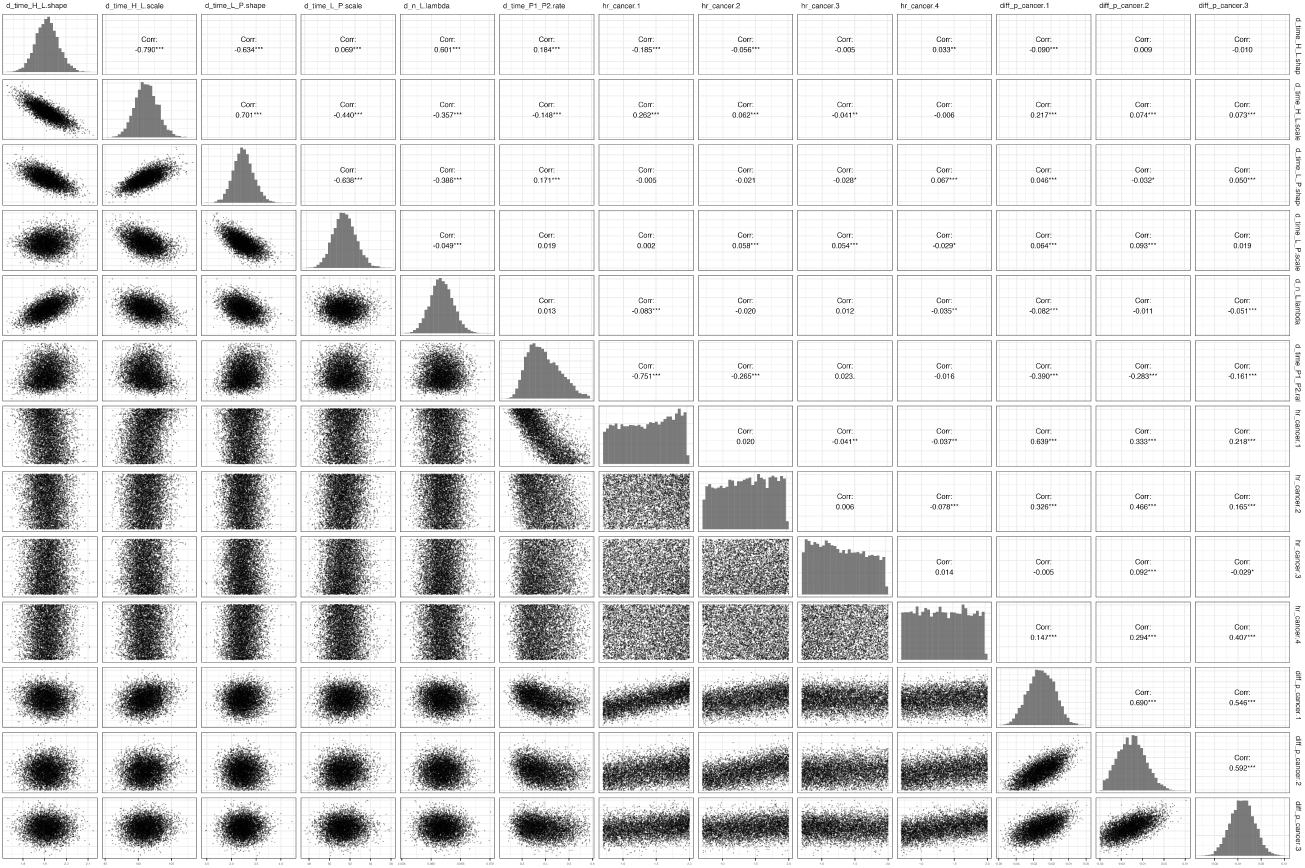
BayCANN correlation plot

